# Pulmonary function in young childhood cancer survivors: results from a prospective multicentre cohort

**DOI:** 10.1101/2025.09.19.25336186

**Authors:** Maša Žarković, Christina Schindera, Nicolas Waespe, Daniel Trachsel, Anne Mornand, Marc Ansari, Philipp Latzin, Claudia E Kuehni

**Affiliations:** Childhood Cancer Research Group, Institute of Social and Preventive Medicine, University of Bern, Switzerland; Graduate School for Health Sciences, University of Bern, Switzerland; Division of Paediatric Oncology/Haematology, University Children’s Hospital Basel, University of Basel, Switzerland; Division of Pediatric Hematology and Oncology, Department of Pediatrics, Inselspital, Bern University Hospital, University of Bern, Switzerland; CANSEARCH Research Platform for Pediatric Oncology and Hematology, Department of Pediatrics, Gynecology and Obstetrics, Faculty of Medicine, University of Geneva, Switzerland; Pediatric Intensive Care and Pulmonology, University Children’s Hospital Basel, Basel, Switzerland; Division of Pediatric Pulmonology, Department of Women, Child and Adolescent, University Hospital of Geneva, Switzerland; Division of Pediatric Oncology and Hematology, Department of Women, Child and Adolescent, University Hospital of Geneva, Switzerland; Division of Paediatric Respiratory Medicine and Allergology, Department of Paediatrics, Inselspital, Bern University Hospital, University of Bern, Switzerland

## Abstract

**Background:** Childhood cancer survivors (CCS) are at risk of pulmonary late effects, but post-treatment lung function remains understudied. Current guidelines recommend screening only for symptomatic survivors treated with lung-damaging treatments. We evaluated pulmonary function, risk factors, and respiratory symptoms in a broad paediatric CCS cohort, including those with standard treatments.

**Methods:** In this prospective multicenter study, we included CCS aged 6–21 years, stratified as high-risk (thoracic radiotherapy/surgery, busulfan/bleomycin/nitrosourea chemotherapy, haematopoietic stem cell transplantation [HSCT]) or standard-risk (other systemic treatment). Pulmonary function was assessed via spirometry (forced expiratory volume in 1 second [FEV₁], forced vital capacity [FVC]), body plethysmography (total lung capacity [TLC]), and diffusing capacity for carbon monoxide (DLCO), expressed as z-scores using Global Lung Initiative references. We assessed respiratory symptoms via questionnaires and analysed treatment associations with pulmonary function using multivariable linear regression.

**Results:** With a response rate of 90%, 251 CCS participated (median 7 years post-diagnosis). Mean z-scores for FEV_1_, FVC, TLC, and DLCO were lower in high-risk (-0.70, -0.91, -0.54, -0.17, respectively) than standard-risk survivors (-0.10, -0.22, -0.17, 0.32). Thoracic surgery and nitrosoureas were associated with lower TLC (-0.53, -1.37), HSCT with reduced FEV_1_ and FVC (-0.80, –0.84), and thoracic radiotherapy with lower DLCO (-0.63). Respiratory symptoms were reported by 32%, but 64% of those with impaired lung function were asymptomatic.

**Conclusion:** Pulmonary function was mostly normal in standard-risk CCS, but impaired in high-risk, although often asymptomatic. These findings support targeted surveillance based on treatment exposure rather than symptoms to guide long-term care.

## Introduction

Survival after childhood cancer has markedly improved in recent decades, with over 80% of patients in high-income countries now expected to become long-term survivors [1–3]. However, this success has brought an increasing burden of late treatment-related complications [4]. Pulmonary morbidity is among the leading causes of non-malignant mortality in childhood cancer survivors (CCS) [5]. Several cancer treatments have been found to adversely affect the lungs, including chemotherapeutic agents such as bleomycin, busulfan, and nitrosoureas, thoracic surgery, radiotherapy, and haematopoietic stem cell transplantation (HSCT). Damage to the lung parenchyma, vasculature, or airways [6,7], may result in restrictive, obstructive, or diffusion impairments on pulmonary function tests (PFTs), with cumulative incidence increasing over time since diagnosis [4].

We recently led the development of the International Late Effects of Childhood Cancer Guideline Harmonisation Group (IGHG) recommendations for pulmonary surveillance in CCS, which were based on a systematic review of the available literature [8]. This process identified considerable methodological weaknesses in existing studies, including predominantly retrospective designs, non-standardised PFT assessments, and inconsistent use of Global Lung Initiative (GLI)-based reference values. Many studies focused on selected subgroups, such as HSCT recipients [9–11] or those treated with thoracic radiotherapy [12,13], while others included only adult survivors [14–16], leaving important gaps in early post-treatment data and risk factor assessment in childhood and adolescence [17]. Evidence was insufficient to assess pulmonary outcomes in survivors treated with other systemic therapies not previously established as lung-damaging, and to evaluate how respiratory symptoms relate to PFT impairments. As a result, the IGHG could only recommend routine PFTs for symptomatic survivors exposed to known lung-damaging treatments.

Therefore, we conducted a prospective cohort study of paediatric CCS with diverse treatment exposures. We aimed to determine the prevalence of pulmonary function abnormalities using standardised PFTs, evaluate associations with clinical and treatment-related factors, and assess the frequency of respiratory symptoms in relation to PFT impairments.

## Methods

### Study design and procedure

The Swiss Childhood Cancer Survivor Study FollowUp–Pulmo (SCCSS-FU Pulmo) is a prospective cohort evaluating long-term pulmonary health in CCS. It is embedded in routine follow-up care at the University Children’s Hospitals of Bern, Basel, and Geneva, and linked to the Swiss Childhood Cancer Registry (ChCR), a nationwide registry that captures over 95% of children diagnosed with cancer [18] according to the International Classification of Childhood Cancer (ICCC-3) [19].

To ensure near-complete recruitment, eligible survivors were identified through a two-way approach: local study teams regularly screened upcoming follow-up appointments and cross-checked against lists from the ChCR to identify any additional eligible patients. The treating physicians approved and scheduled PFTs as part of routine oncological care. Patients received an information letter, consent form, and respiratory questionnaire before the visit. If consent and questionnaire were not returned before or during the visit, up to two reminders were sent to allow inclusion of data. Detailed procedures have been published [20]. The study was approved by the Ethics Committee of the Canton of Bern (KEK-BE: 2019-00739).

### Study population

We included all CCS aged 6 to 21 years who had completed treatment, were in remission, and still in follow-up care at participating centers between June 2022 and July 2025. Survivors treated only with surgery or radiotherapy outside the thorax were excluded due to minimal expected impact on pulmonary function.

Participants were classified into two risk groups. High-risk survivors had received therapies previously associated with impaired pulmonary outcomes, including pulmotoxic chemotherapy (busulfan, bleomycin, carmustine or lomustine), pulmotoxic radiotherapy (radiation to the chest (mantle, mediastinal, or whole lung fields), abdomen (whole or any upper field), or total body irradiation), HSCT, or thoracic surgery (any procedure involving the lungs or chest wall such as lobectomy, wedge resection, or thoracotomy) [6,21]. Standard-risk survivors were treated with any other systemic anticancer therapy (chemotherapy, immunotherapy, or targeted agents).

In a secondary step, we further subdivided the high-risk group by thoracic surgery to distinguish the potential effects of surgical interventions from those of other high-risk treatments.

### Pulmonary function tests

Pulmonary function assessment included spirometry, body plethysmography, and diffusing capacity of the lung for carbon monoxide (DLCO), performed according to European Respiratory Society/American Thoracic Society (ERS/ATS) guidelines [22–24]. Tests were conducted in paediatric lung function laboratories by trained technicians with on-site quality control, and results were validated by paediatric pulmonologists. Pulmonary function outcomes were grouped into three physiological domains: markers of restriction (forced vital capacity [FVC], total lung capacity [TLC], and functional residual capacity [FRC]); markers of obstruction (forced expiratory volume in 1 second [FEV_1_], FEV_1_/FVC ratio, and forced mid-expiratory flow [FEF_25–75%_]); and markers of diffusion capacity (DLCO and carbon monoxide transfer coefficient [KCO]). Spirometry was performed pre-bronchodilation, and DLCO values were corrected for haemoglobin concentration if a measurement was available within 30 days. Outcomes were expressed as sex-, age-, height-, and ethnicity-adjusted z-scores using GLI reference equations, calculated with the official GLI calculator (https://gli-calculator.ersnet.org/) [25–28]. Lower limit of normal (LLN) was defined as z-score < -1.645. Further details on parameters and reference standards are provided in Supplementary Tables S1 and S2.

### Respiratory symptoms

At the time of pulmonary function assessment, participants completed a questionnaire assessing respiratory symptoms in the past 12 months. Questionnaires were parent-completed for children <14 years and self-completed by older participants. The questionnaire included items on respiratory symptoms at rest and during exertion (wheeze, dyspnea, and cough), and chronic cough (>4 weeks). English translations of original German/French questions are provided in Supplementary Table S1.

### Clinical and demographic characteristics

We extracted clinical and demographic information from the medical records of participating centres. This included sex, age at study, age at cancer diagnosis, time since diagnosis, cancer treatments, and anthropometric measures (height, weight, and body mass index), converted into z-scores using World Health Organisation references [29]. Current asthma was defined based on either a documented diagnosis in medical records or self-report in the questionnaire.

### Statistical analysis

We summarised continuous variables using means with 95% confidence intervals (CI) or medians with interquartile ranges (IQR), and categorical variables as frequencies with percentages. In the first analytical step, we compared PFT outcomes between the standard-risk and high-risk groups using independent two-sample t-tests. In a second step, we further stratified the high-risk group by thoracic surgery and compared PFT outcomes between the standard-risk group and the two high-risk subgroups using t-tests to assess whether results (e.g. restriction) were explained by thoracic surgery alone. Categorical variables were compared using chi-square or Fisher’s exact test.

In addition to comparing risk groups, we also examined associations with individual treatment exposures using multivariable linear regression models. Exposures were selected a priori based on clinical relevance and previous evidence of pulmonary effects. For each physiological domain, we used one key outcome as the dependent variable: FEV_1_/FVC z-ratio (obstruction), TLC z-score (restriction), and DLCO z-score (diffusion). Exposure variables included pulmotoxic chemotherapy agents (bleomycin, busulfan, nitrosoureas), thoracic surgery, pulmotoxic radiotherapy, and HSCT. All models were adjusted for age at study, sex, and time since diagnosis. In sensitivity analyses, we additionally adjusted for asthma, to account for potential confounding by pre-existing or concurrent respiratory disease, and for study center, to assess whether differences between centers affected the results. We used Stata version 16.1 (StataCorp LLC) and R version 4.4.2 (R Foundation for Statistical Computing).

## Results

### Characteristics of the study population

Of 278 eligible survivors identified across participating centers, 251 participated (response rate 90%) (Supplementary Figure S1). The median age at study was 14 years (IQR 10–17) and median time since diagnosis was 7 years (IQR 4–9) (Table 1). The most common cancer diagnoses were leukemia (49%), lymphoma (12%), and neuroblastoma (9%). Almost all participants (99%) received chemotherapy, including 28 (11%) with pulmotoxic chemotherapy. Thirty-five (14%) received pulmotoxic radiotherapy, 20 (8%) underwent thoracic surgery, and 33 (13%) HSCT.

**Table 1.**
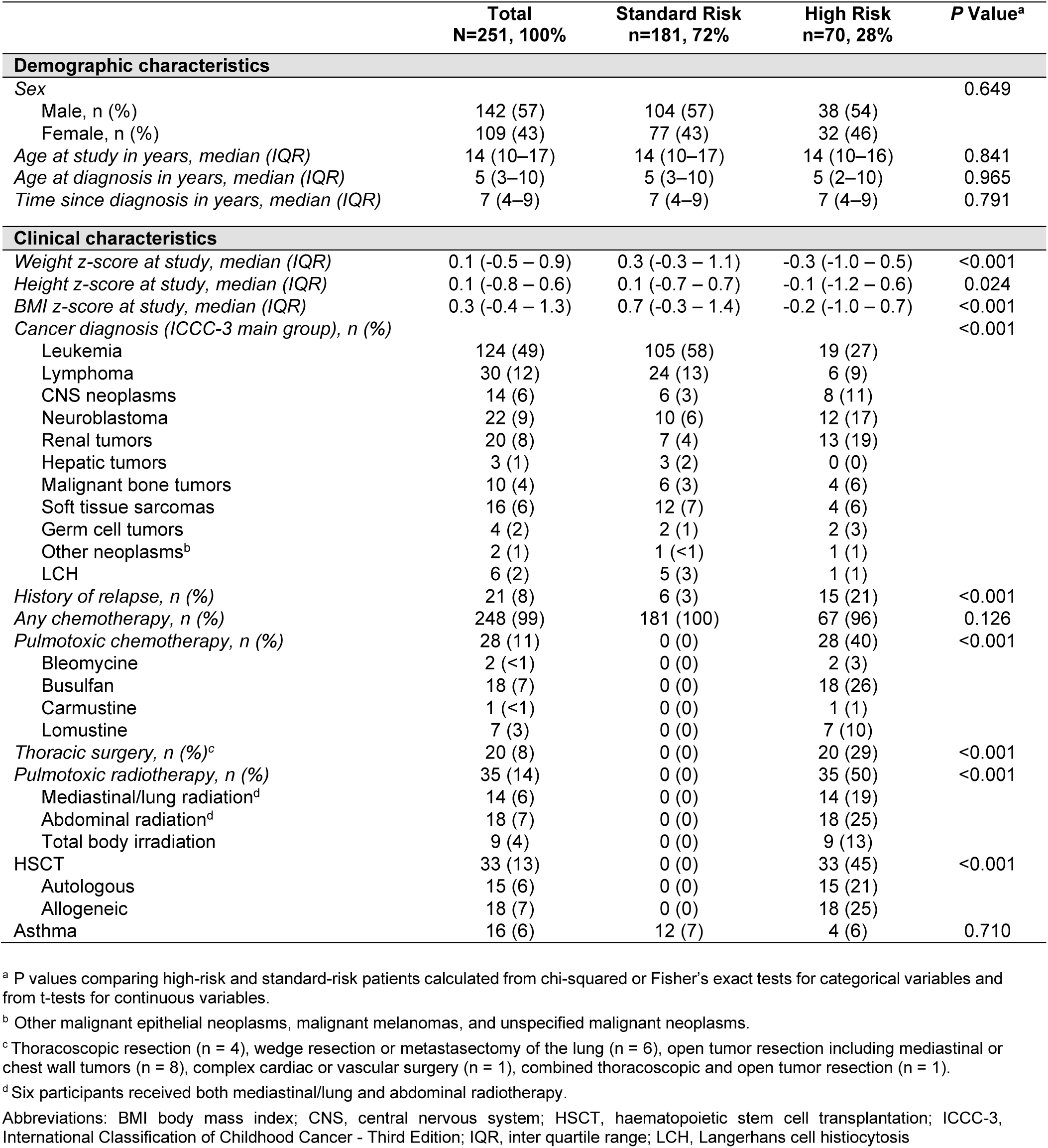
Characteristics of participating childhood cancer survivors, overall and stratified by treatment-related risk groups.

Standard-risk survivors accounted for 72% of the cohort, while 28% were classified as high-risk. High-risk survivors were shorter (median z-score –0.1 vs. 0.1), slimmer (-0.3 vs. 0.3), and had lower BMI z-scores (–0.2 vs. 0.7) than standard-risk. Cancer types differed by risk group, reflecting distinct treatment protocols. A current asthma diagnosis was found in 16 (6%).

### Pulmonary function parameters

PFTs were successful in most of the 251 participants: 95% had valid spirometry, 86% body plethysmography, and 79% DLCO, with lower completion rates for the latter two due to limited cooperation in younger children (Supplementary Figure S1, Table 2).

**Table 2.**
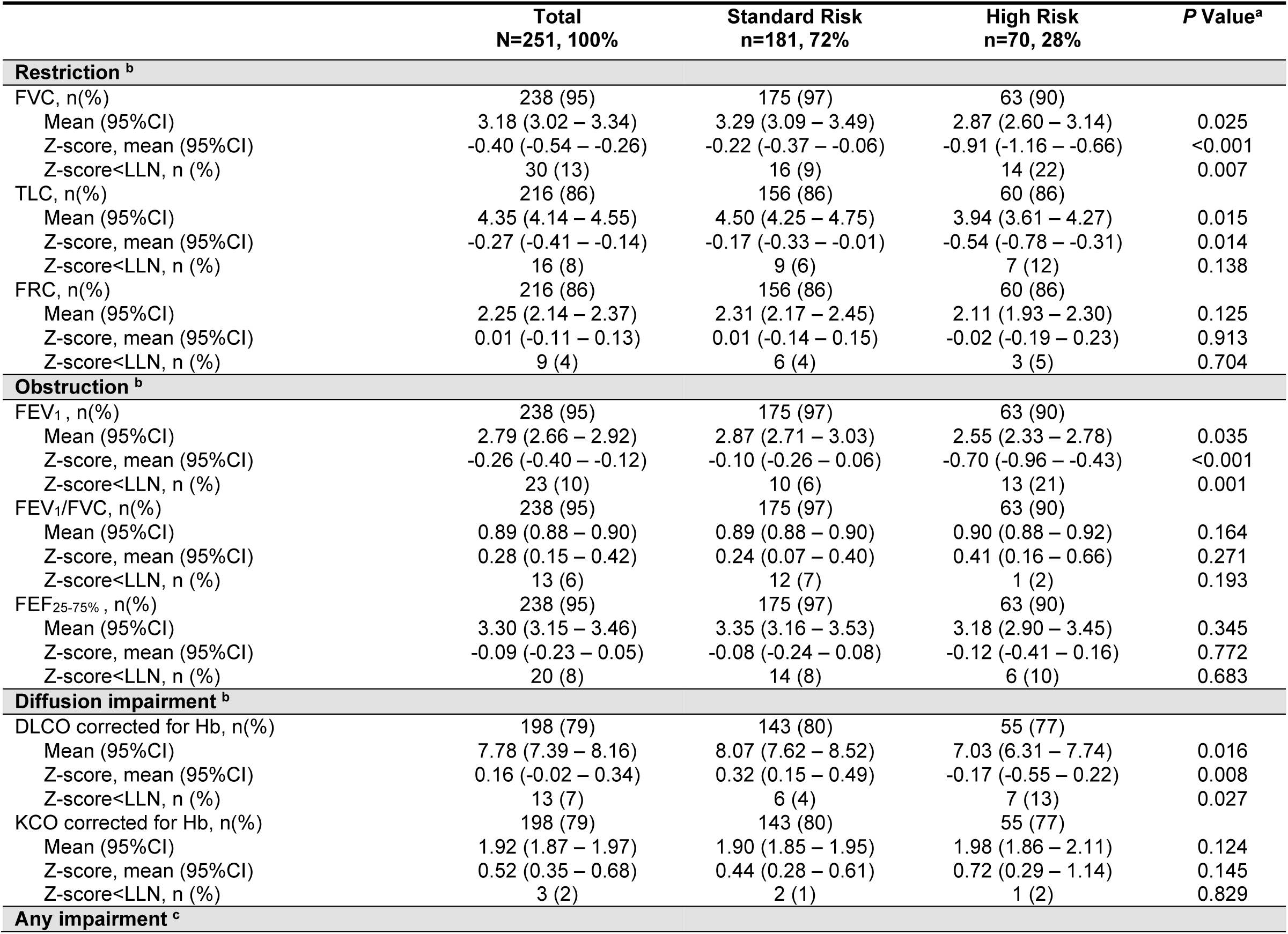
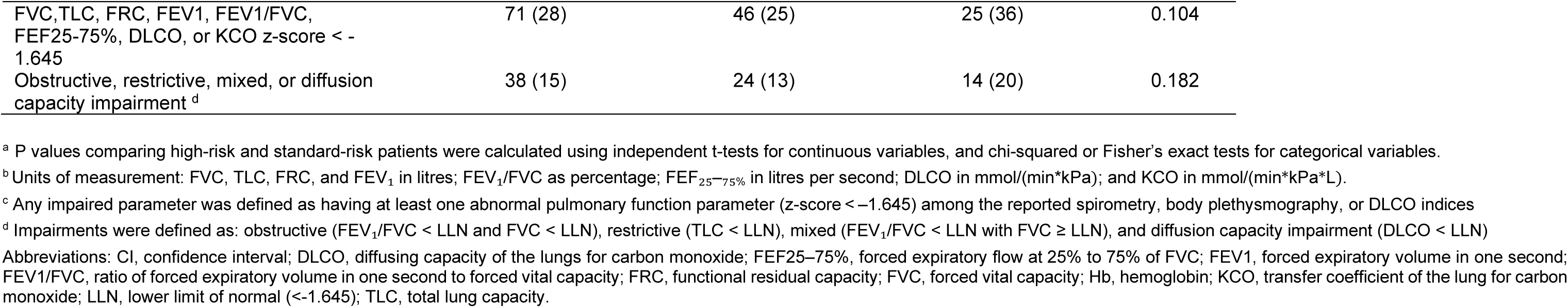
Pulmonary function parameters among childhood cancer survivors, reported overall and stratified by treatment-related risk groups. Parameters include measures of restriction, obstruction, and diffusion capacity, with prevalence of impairment and group comparisons.

|  | Total<br>N=251, 100% | Standard Risk<br>n=181, 72% | High Risk<br>n=70, 28% | P Value<sup>a</sup> |
| --- | --- | --- | --- | --- |
| **Restriction<sup>b</sup>** |  |  |  |  |
| FVC, n(%) | 238 (95) | 175 (97) | 63 (90) |  |
| Mean (95%CI) | 3.18 (3.02 – 3.34) | 3.29 (3.09 – 3.49) | 2.87 (2.60 – 3.14) | 0.025 |
| Z-score, mean (95%CI) | -0.40 (-0.54 – -0.26) | -0.22 (-0.37 – -0.06) | -0.91 (-1.16 – -0.66) | <0.001 |
| Z-score<LLN, n (%) | 30 (13) | 16 (9) | 14 (22) | 0.007 |
| TLC, n(%) | 216 (86) | 156 (86) | 60 (86) |  |
| Mean (95%CI) | 4.35 (4.14 – 4.55) | 4.50 (4.25 – 4.75) | 3.94 (3.61 – 4.27) | 0.015 |
| Z-score, mean (95%CI) | -0.27 (-0.41 – -0.14) | -0.17 (-0.33 – -0.01) | -0.54 (-0.78 – -0.31) | 0.014 |
| Z-score<LLN, n (%) | 16 (8) | 9 (6) | 7 (12) | 0.138 |
| FRC, n(%) | 216 (86) | 156 (86) | 60 (86) |  |
| Mean (95%CI) | 2.25 (2.14 – 2.37) | 2.31 (2.17 – 2.45) | 2.11 (1.93 – 2.30) | 0.125 |
| Z-score, mean (95%CI) | 0.01 (-0.11 – 0.13) | 0.01 (-0.14 – 0.15) | -0.02 (-0.19 – 0.23) | 0.913 |
| Z-score<LLN, n (%) | 9 (4) | 6 (4) | 3 (5) | 0.704 |
| **Obstruction<sup>b</sup>** |  |  |  |  |
| FEV<sub>1</sub>, n(%) | 238 (95) | 175 (97) | 63 (90) |  |
| Mean (95%CI) | 2.79 (2.66 – 2.92) | 2.87 (2.71 – 3.03) | 2.55 (2.33 – 2.78) | 0.035 |
| Z-score, mean (95%CI) | -0.26 (-0.40 – -0.12) | -0.10 (-0.26 – 0.06) | -0.70 (-0.96 – -0.43) | <0.001 |
| Z-score<LLN, n (%) | 23 (10) | 10 (6) | 13 (21) | 0.001 |
| FEV<sub>1</sub>/FVC, n(%) | 238 (95) | 175 (97) | 63 (90) |  |
| Mean (95%CI) | 0.89 (0.88 – 0.90) | 0.89 (0.88 – 0.90) | 0.90 (0.88 – 0.92) | 0.164 |
| Z-score, mean (95%CI) | 0.28 (0.15 – 0.42) | 0.24 (0.07 – 0.40) | 0.41 (0.16 – 0.66) | 0.271 |
| Z-score<LLN, n (%) | 13 (6) | 12 (7) | 1 (2) | 0.193 |
| FEF<sub>25-75%</sub>, n(%) | 238 (95) | 175 (97) | 63 (90) |  |
| Mean (95%CI) | 3.30 (3.15 – 3.46) | 3.35 (3.16 – 3.53) | 3.18 (2.90 – 3.45) | 0.345 |
| Z-score, mean (95%CI) | -0.09 (-0.23 – 0.05) | -0.08 (-0.24 – 0.08) | -0.12 (-0.41 – 0.16) | 0.772 |
| Z-score<LLN, n (%) | 20 (8) | 14 (8) | 6 (10) | 0.683 |
| **Diffusion impairment<sup>b</sup>** |  |  |  |  |
| DLCO corrected for Hb, n(%) | 198 (79) | 143 (80) | 55 (77) |  |
| Mean (95%CI) | 7.78 (7.39 – 8.16) | 8.07 (7.62 – 8.52) | 7.03 (6.31 – 7.74) | 0.016 |
| Z-score, mean (95%CI) | 0.16 (-0.02 – 0.34) | 0.32 (0.15 – 0.49) | -0.17 (-0.55 – 0.22) | 0.008 |
| Z-score<LLN, n (%) | 13 (7) | 6 (4) | 7 (13) | 0.027 |
| KCO corrected for Hb, n(%) | 198 (79) | 143 (80) | 55 (77) |  |
| Mean (95%CI) | 1.92 (1.87 – 1.97) | 1.90 (1.85 – 1.95) | 1.98 (1.86 – 2.11) | 0.124 |
| Z-score, mean (95%CI) | 0.52 (0.35 – 0.68) | 0.44 (0.28 – 0.61) | 0.72 (0.29 – 1.14) | 0.145 |
| Z-score<LLN, n (%) | 3 (2) | 2 (1) | 1 (2) | 0.829 |
| **Any impairment<sup>c</sup>** |  |  |  |  |
| FVC, TLC, FRC, FEV<sub>1</sub>, FEV<sub>1</sub>/FVC, FEF<sub>25-75%</sub>, DLCO, or KCO z-score < -1.645 | 71 (28) | 46 (25) | 25 (36) | 0.104 |
| Obstructive, restrictive, mixed, or diffusion capacity impairment<sup>d</sup> | 38 (15) | 24 (13) | 14 (20) | 0.182 |

<sup>a</sup> P values comparing high-risk and standard-risk patients were calculated using independent t-tests for continuous variables, and chi-squared or Fisher's exact tests for categorical variables.
<sup>b</sup> Units of measurement: FVC, TLC, FRC, and FEV<sub>1</sub> in litres; FEV<sub>1</sub>/FVC as percentage; FEF<sub>25-75%</sub> in litres per second; DLCO in mmol/(min\*kPa); and KCO in mmol/(min\*kPa\*L).
<sup>c</sup> Any impaired parameter was defined as having at least one abnormal pulmonary function parameter (z-score < -1.645) among the reported spirometry, body plethysmography, or DLCO indices
<sup>d</sup> Impairments were defined as: obstructive (FEV<sub>1</sub>/FVC < LLN and FVC < LLN), restrictive (TLC < LLN), mixed (FEV<sub>1</sub>/FVC < LLN with FVC ≥ LLN), and diffusion capacity impairment (DLCO < LLN)
Abbreviations: CI, confidence interval; DLCO, diffusing capacity of the lungs for carbon monoxide; FEF<sub>25-75%</sub>, forced expiratory flow at 25% to 75% of FVC; FEV<sub>1</sub>, forced expiratory volume in one second; FEV<sub>1</sub>/FVC, ratio of forced expiratory volume in one second to forced vital capacity; FRC, functional residual capacity; FVC, forced vital capacity; Hb, hemoglobin; KCO, transfer coefficient of the lung for carbon monoxide; LLN, lower limit of normal (<-1.645); TLC, total lung capacity.

#### Parameters indicating possible restriction

The mean FVC z-score was -0.22 (95%CI -0.37 – -0.06) in standard-risk, only slightly below the population mean (z-score=0), while in high-risk survivors it was even lower (-0.91 [-1.16 – - 0.66]). TLC showed a similar pattern, with z-scores of -0.17 (-0.33 – -0.01) in the standard-risk and -0.54 (-0.78 – -0.31) in the high-risk group. Compared to standard-risk, high-risk participants had lower FVC (*p* < 0.001) and TLC (*p* = 0.014). Restrictive impairment (TLC < LLN) [30] was present in 8%, more frequently in the high-risk (12%) than the standard-risk group (6%).

#### Parameters indicating possible obstruction

The mean FEV₁ z-score was within normal limits in the standard-risk (-0.10 [-0.26 to 0.06]), but reduced in the high-risk group (-0.70 [-0.96 – -0.43]). The FEV₁/FVC ratio remained preserved in both groups (z-scores: 0.24 and 0.41), and FEF_25-75%_ did not differ. Obstructive impairment (FEV₁/FVC < LLN with FVC ≥ LLN) [30] was rare (4% overall), occurring slightly more often in the standard-risk group (6% vs. 0%). Mixed impairment (FEV₁/FVC < LLN and FVC < LLN) [30] was found in only 2 (1%) participants.

#### Diffusion capacity parameters

DLCO was normal in the standard-risk group (mean z-score 0.32 [0.15 – 0.49]), while the high-risk group showed slightly reduced values (-0.27 [-0.72 – 0.18]). Diffusion impairment (DLCO < LLN) [30] was found in 13% of high-risk compared to 4% of standard-risk survivors. KCO did not differ between groups and was impaired in only 2%.

Further stratification of the high-risk group based on thoracic surgery showed that restrictive changes were similarly frequent in both risk groups (12% and 11%), but TLC was more reduced after surgery (z-score -0.73 vs -0.45). Diffusion impairment was confined to those without thoracic surgery but with other high-risk treatments (18% vs. 0%), reflected by lower mean DLCO z-scores (-0.23 vs. -0.01) (Supplementary Table S3, Figure 1).

**Figure 1.**
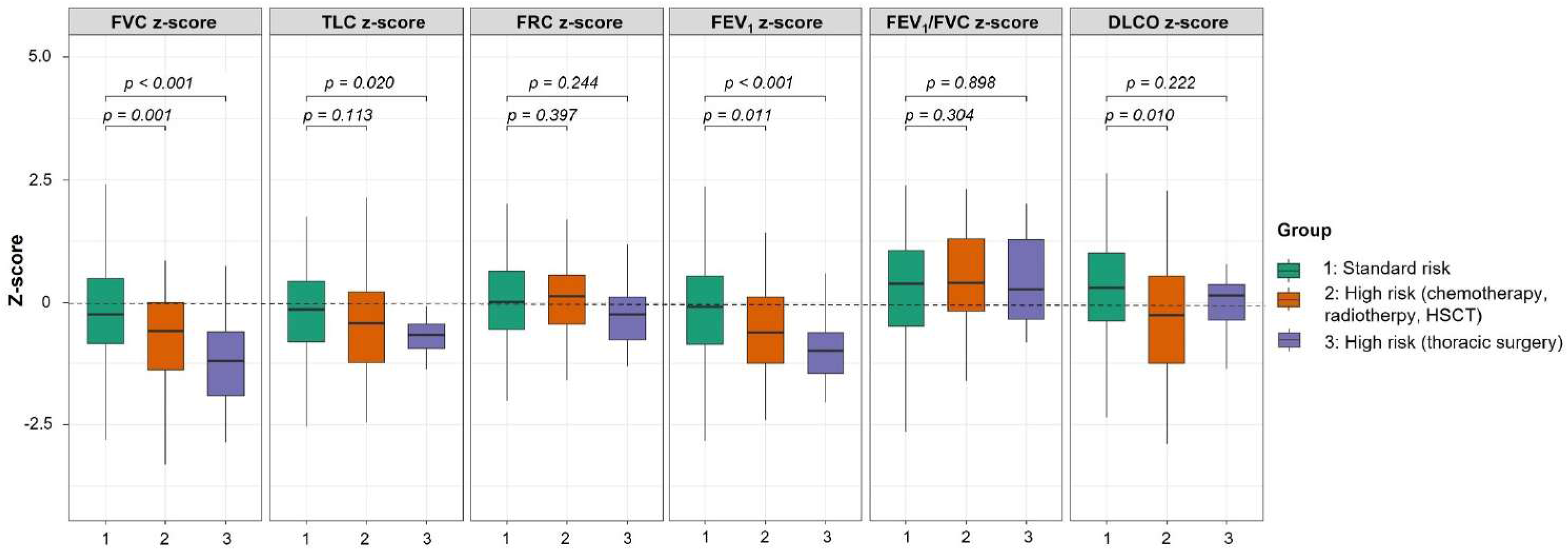
Pulmonary function outcomes in childhood cancer survivors compared across risk groups: standard risk (exposed to any systemic anticancer treatment), high risk (exposed to pulmotoxic chemotherapy, radiotherapy, or HSCT, or combinations of those treatments) and high risk who underwent thoracic surgery. Box plots show median z-scores and interquartile ranges (coloured), with the dashed line indicating z-score = 0. P-values were calculated using two-sided independent t-tests comparing standard risk vs high-risk groups. Abbreviations: _DLCO, diffusing capacity of the lungs for carbon monoxide; FEV1, forced expiratory volume in one second; FEV1/FVC, ratio of forced expiratory volume in one second to forced vital capacity; FRC, functional residual capacity; FVC, forced vital capacity; HSCT, haematopoietic stem cell transplantation; TLC, total lung capacity._

### Association between treatment exposures and pulmonary function

In multivariable models, several treatment exposures were associated with impaired pulmonary function (Table 3). Nitrosoureas were linked to reduced TLC z-scores (-1.37 [-2.18 – -0.57]) and lower FEV_1_/FVC ratios (0.05 [0.01 – 0.10]). Thoracic surgery was associated with reduced TLC z-scores (-0.53 [-1.01 – -0.05]), while pulmotoxic radiotherapy with lower DLCO z-scores (-0.63 [-1.27 – -0.01]). Nitrosoureas, thoracic surgery, and HSCT were associated with both reduced FEV_1_ and FVC z-scores (Supplementary Table S4).

**Table 3.**
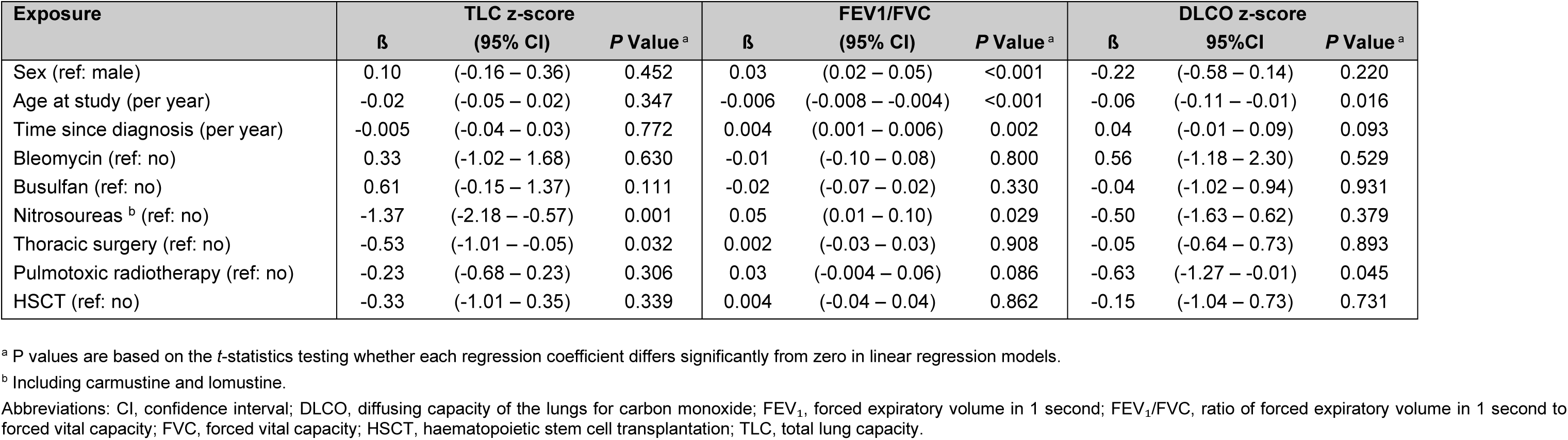
Predictors of pulmonary function outcomes among childhood cancer survivors. Multivariable linear regression models showing associations of clinical characteristics and cancer treatments with indicators of obstruction (FEV1/FVC), restriction (TLC z-score), and diffusion impairment (DLCO z-score).

Among clinical covariates, older age at study was associated with lower FEV_1_, FVC, and DLCO z-scores. In sensitivity analyses, additionally adjusting for asthma, results remained similar (Supplementary Table S5); adjustment for study center did not change the effect estimates. Asthma was associated with lower FEV_1_ and FEV_1_/FVC.

### Respiratory symptoms and their association with pulmonary function

Thirty-two percent (n=81) reported at least one respiratory symptom within the previous 12 months, with similar rates in standard- and high-risk groups (31% vs. 34%; Supplementary Table S6).

We defined PFT impairment as any parameter (FEV_1_, FVC, FEV_1_/FVC, FEF_25-75%_, TLC, FRC, DLCO, or KCO) below the LLN. Among those with impairment (n = 69), 36% reported at least one symptom, while 64% were asymptomatic. This pattern was similar across risk groups: 56% of high-risk and 67% of standard-risk survivors with impairment had no symptoms. Of those with symptoms (n = 81), 31% had PFT impairment. There was no clear association between symptoms and PFT impairment (Figure 2, Supplementary Table S7).

**Figure 2.**
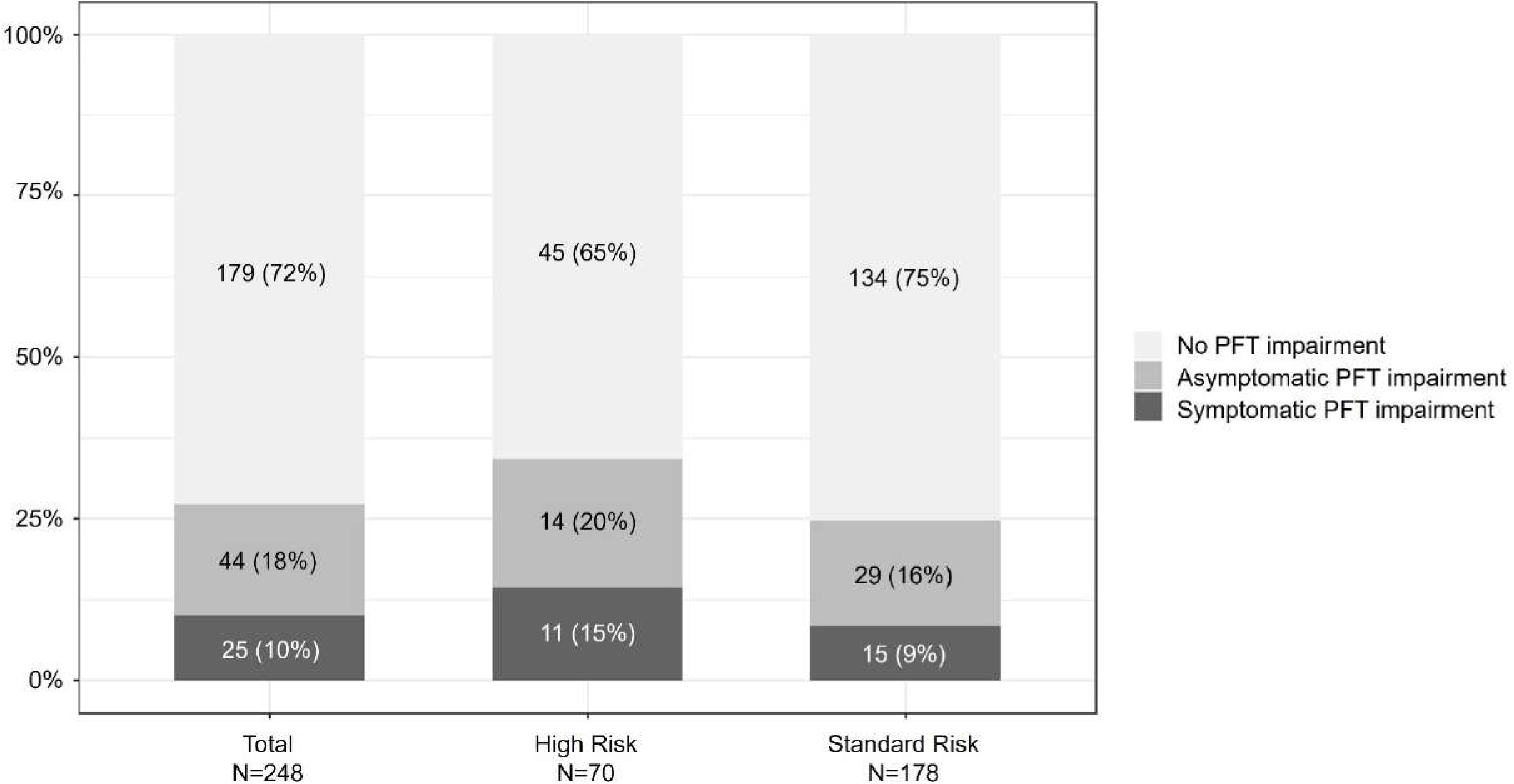
Proportion of childhood cancer survivors with no, asymptomatic, and symptomatic pulmonary function test impairment, stratified by treatment-related risk groups. The bars represent total population, high risk and standard risk groups, excluding three participants without questionnaire data. Pulmonary function test (PFT) impairment was defined as having at least one pathological z-score value of either FEV1, FVC, FEV1/FVC, FEF25-75%, TLC, FRC, DLCO, KCO below the lower limit of normal (z-score <-1.645). The white areas represent the proportion of participants with PFT impairment but no reported symptoms. Reported symptoms were considered as occurring in the past 12 months and having had at least one of the following symptoms: at rest (wheeze, dyspnea, cough), during exertion (wheeze, dyspnea, cough), or chronic cough lasting for more than 4 weeks. Grey areas represent participants who didn’t report any of the symptoms but had a PFT impairment, while black areas represent participants who reported symptoms and had a PFT impairment. Abbreviations: DLCO, diffusing capacity of the lungs for carbon monoxide; FEF25–75%, forced expiratory flow at 25% to 75% of FVC; FEV1, forced expiratory volume in one second; FEV1/FVC, ratio of forced expiratory volume in one second to forced vital capacity; FRC, functional residual capacity; FVC, forced vital capacity; KCO, transfer coefficient of the lung for carbon monoxide; TLC, total lung capacity.

## Discussion

In this prospective cohort study of CCS early (median 7 years) after cancer diagnosis, pulmonary function test abnormalities were uncommon. Lung function in the standard-risk group was largely within normal ranges, while high-risk survivors more frequently showed restrictive and diffusion impairments. Nitrosoureas, thoracic surgery, and HSCT were associated with abnormalities suggestive of restrictive or non-specific ventilatory defects, while radiotherapy was linked to diffusion impairments. One-third reported respiratory symptoms, but over half of those with impaired pulmonary function remained asymptomatic.

Restrictive and diffusion impairments were the most common patterns, affecting 8% and 7% of survivors, and occurred more frequently in high-risk survivors (12% and 13%). These rates exceed the 5% expected in a healthy population (defining LLN as -1.645 z-scores) and may indicate effects of lung-damaging treatments. Comparisons with prior studies are limited by inconsistent definitions, with many using %predicted thresholds not standardised for age, sex, and height (Supplementary Table S8). We previously reported higher GLI z-score-based prevalences from Switzerland. A retrospective nationwide study of survivors with the same high-risk profile, but on average 10 years post-treatment, found a prevalence of 22% restriction and 25% diffusion impairment [31], likely reflecting selection bias, as lung function data may have been more available for symptomatic CCS or those treated more intensively. Similarly, in a retrospective study of CCS post-HSCT, we found 32% restriction [32]. Even higher rates have been observed in adult high-risk survivors in the Netherlands (34% restriction, 42% diffusion impairment) [33]. The lower prevalence observed here may reflect a shorter time since diagnosis. However, not all observed patterns necessarily reflect intrinsic pulmonary pathology, as reduced DLCO with preserved or increased KCO can indicate incomplete alveolar expansion [34], and reduced TLC with preserved FRC can suggest reduced thoracic expansion rather than parenchymal disease [30].

Obstructive abnormalities were within the normal range expected for the general population (4% overall, 1% in the high-risk group), consistent with findings from young CCS cohorts (median age 16 years) showing 1–2% obstruction [31,32]. Rates appear to remain low even with longer follow-up, such as 7% in our previous study of an adult high-risk CCS cohort (median age 30 years) [35]. These findings support that obstruction is not a predominant pattern in CCS. The standard-risk group showed overall normal lung function, with <6% impairments for each outcome. With most studies focusing on high-risk groups, only a few in leukemia survivors allow relevant comparison. In a 1998 Danish study (median age 16.2 years) 8% FEV₁, 15% FVC, and 11% TLC were below z-score -1.645 [36,37]. Another study in leukemia survivors (median age 14.6 years) using <80%predicted thresholds found 20–23% impairments for FEV_1_, FVC, TLC, DLCO. These higher rates may reflect older, more toxic treatment regimens [37]. Mean FEV_1_ and FVC z-scores in our standard-risk group (-0.10 and -0.22) were slightly higher than those reported in a Swiss general paediatric population (-0.50 and -0.43), indicating near-normal lung function and potential limitations of GLI reference values for Swiss children [38].

Several cancer treatments, including thoracic surgery, radiotherapy, HSCT, and nitrosoureas were associated with reduced pulmonary function, and associations persisted after adjusting for asthma. Thoracic surgery was associated with the most pronounced decreases in FEV_1_, FVC, and TLC, likely reflecting reduced lung volume or chest wall compliance, as previously reported in survivors undergoing metastasectomy or structural resections [15,39,40]. Radiotherapy was associated with DLCO impairment, but not with reduced TLC in our cohort. This differs from findings in adult survivors where both parameters were often affected [14,15], and may reflect the shorter latency in our younger population. Nitrosoureas were independently associated with reduced TLC and higher FEV_1_/FVC, suggesting restrictive changes. These agents, known for their alkylating and carbamylating properties, have been implicated in delayed-onset pulmonary toxicity through interstitial damage and inflammation [7,41]. Similarly, survivors who had undergone HSCT had lower FEV_1_ and FVC, with preserved FEV_1_/FVC and no clear restriction, suggesting a non-specific ventilatory defect possibly related to subtle parenchymal or chest wall changes [10,42]. These findings highlight the importance of considering cumulative exposure profiles in surveillance, as patients with multiple risk factors may be especially vulnerable. Importantly, the expected association of obstructive impairments with asthma supports internal validity [43]. The association of older age at follow-up with lower FEV_1_, FVC, and DLCO may reflect cumulative damage or age-related decline.

Current IGHG guidelines recommend pulmonary function testing in symptomatic CCS, particularly those exposed to thoracic radiotherapy, surgery, or allogeneic HSCT. This approach reflects limited and inconsistent evidence linking treatment exposures to long-term pulmonary dysfunction in asymptomatic survivors. However, our findings suggest that relying on symptoms alone may miss a substantial proportion of cases. Among survivors with impaired pulmonary function, 64% were asymptomatic, a pattern consistent across risk groups and highlighting the frequency of subclinical dysfunction in this population. This may reflect the large pulmonary reserve and the tendency of children and adolescents to underperceive or inadequately communicate respiratory symptoms. Conversely, 32% of survivors reported respiratory symptoms, yet 69% of these had normal PFTs. Such symptoms may also result from transient conditions such as mild infections or allergies, deconditioning, or cardiac conditions. Our results thus underscore the importance of risk-based assessments informed by treatment exposures, rather than symptoms alone, for identifying survivors who may benefit from pulmonary surveillance.

This study is strengthened by a prospective design with a high participation rate (90%), minimising selection bias and supporting the representativeness of our findings. The inclusion of a broad cohort of standard risk survivors provided comprehensive lung function data, which was missing in most previous studies. Reporting pulmonary function as z-scores, rather than %predicted or categorical outcomes, accounts for age-, sex-, and height-related variation, reduces misclassification in paediatric populations, and enables more accurate comparisons across studies and time points. Several limitations should be addressed. First, as in most survivorship studies, pretreatment lung function data were not available. This limits our ability to distinguish treatment-related pulmonary dysfunction from preexisting conditions. However, the median age at diagnosis was 5 years, and reliable results for PFTs are rarely feasible before this age due to insufficient cooperation and test complexity. Second, we lacked detailed data on radiotherapy dosimetry and lung volumes irradiated, which may have led to misclassification of pulmonary radiation exposure severity. Third, the number of survivors exposed to certain treatments was low, which limited statistical power to detect more nuanced associations. To better capture long-term changes, we plan repeated pulmonary assessments over time through ongoing longitudinal follow-up and international pooling of data across prospective cohorts.

In this prospective cohort, lung function in standard-risk CCS was largely within normal limits up to seven years post-treatment, with little evidence of impairments that would currently justify routine screening. In contrast, high-risk survivors, particularly those treated with nitrosoureas, thoracic surgery, HSCT, or pulmotoxic radiotherapy, showed reduced pulmonary function that often remained asymptomatic. These results highlight the importance of treatment-exposure-based monitoring, with post-treatment lung function testing offering early identification of subclinical changes and a reference point for long-term care.

## Data Availability

All data produced in the present study are available upon reasonable request to the authors.

## Statements & Declarations

## Acknowledgements

We thank all survivors for participating in our study, Pediatric Hematology/Oncology and Pediatric Pulmonology teams at the University Hospitals in Bern, Basel, and Geneva, especially the study nurses Sandra Lüscher, Artemis Ionnaki, and Valentine Pradet. We also thank the study team of the Childhood Cancer Research Group and the Childhood Cancer Registry.

This paper incorporated suggestions from AI tools, including ChatGPT (OpenAI) and Grammarly, which were used to improve the clarity, grammar, and readability of the written text. All scientific content, interpretation, and conclusions are the responsibility of the authors.

## Author contribution

Maša Žarković: formal analysis, writing—original draft preparation, visualization; Christina Schindera: writing—review and editing; Nicolas Waespe: writing—review and editing; Daniel Trachsel: writing—review and editing; Anne Mornand: writing—review and editing; Marc Ansari: writing—review and editing; Philipp Latzin: writing—review and editing; Claudia E Kuehni: conceptualization, methodology, writing— review and editing, funding acquisition, supervision.

## Ethics approval

The Ethics Committee of the Canton of Bern (2019-00739) granted ethical approval.

---

**Supplementary Figure S1.**
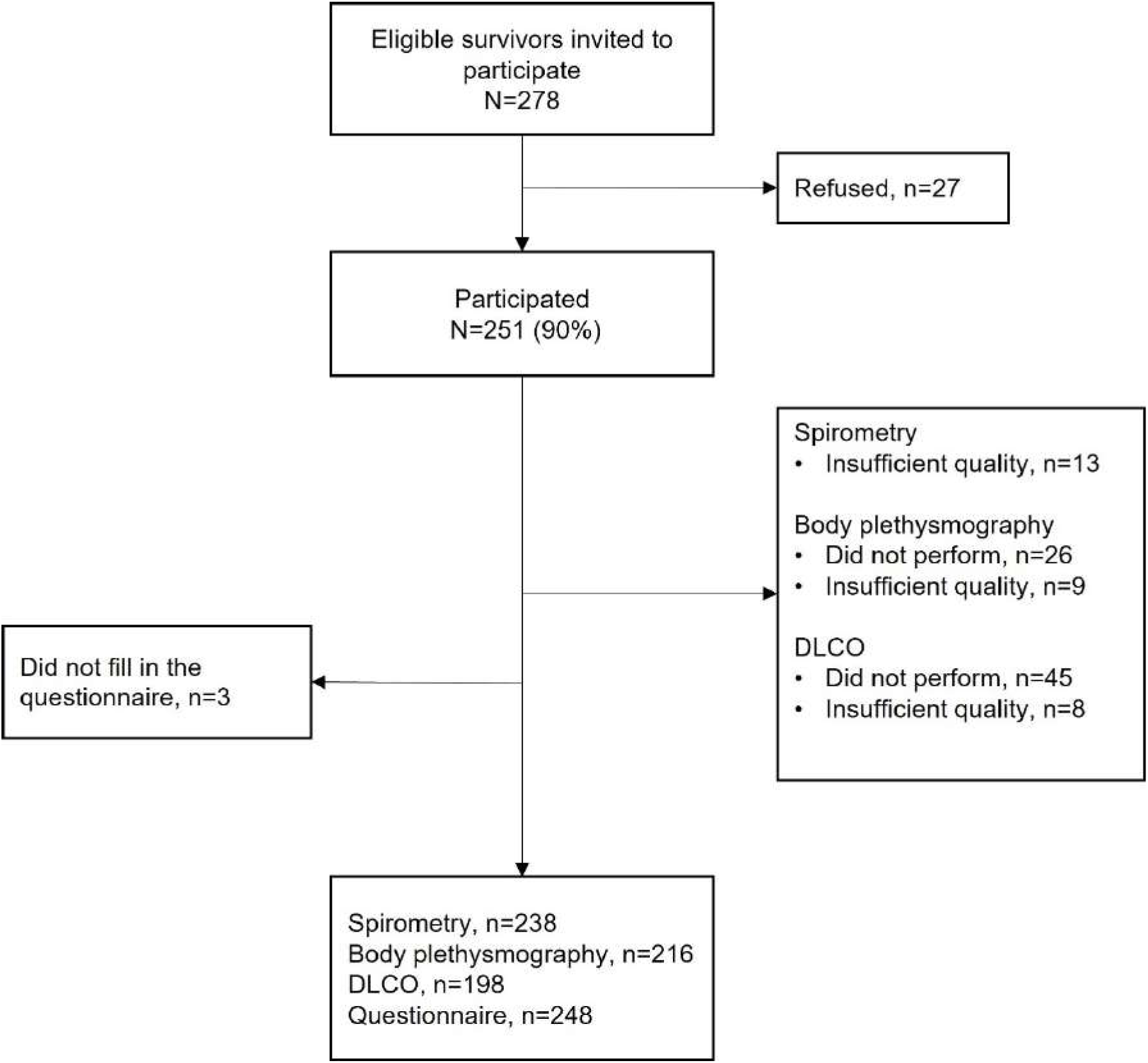
Population tree of study participants of the SCCSS FollowUp – Pulmo study underwent a pulmonary function assessment. All participants underwent spirometry, while some were unable to complete body plethysmography and/or DLCO, primarily due to limited cooperation at younger ages. Abbreviations: DLCO, diffusing capacity of the lungs for carbon monoxide

**Supplementary Table S1.** Pulmonary function tests with main outcome parameters and questions on respiratory symptoms.

| Pulmonary function tests | Unit | Details |
| --- | --- | --- |
| **Spirometry** |  |  |
| FEV<sub>1</sub> | L | Forced expiratory volume in 1 second |
| FVC | L | Forced vital capacity |
| FEV<sub>1</sub>/FVC | % | Ratio of forced expiratory volume in 1 second to forced vital capacity |
| FEF 25-75% | L/s | Forced expiratory flow at 25% to 75% of FVC |
| **Body plethysmography** |  |  |
| TLC | L | Total lung capacity |
| FRC | L | Functional residual capacity |
| **DLCO** |  |  |
| DLCO | mmol/(min*kPa) | Diffusing capacity of the lungs for carbon monoxide |
| KCO | mmol/(min*kPa*L) | Transfer coefficient of the lung for carbon monoxide (calculated as DLCO divided by alveolar volume [VA]) |
| Questionnaire variable | Question |  |
| Respiratory symptoms at rest | Does your child / Do you sometimes have breathing difficulties without exertion? (yes/no)<br>If yes, which? <sup>a</sup> |  |
|  | • Wheeze<br>• Dyspnea<br>• Cough |  |
| Respiratory symptoms during exertion | Does your child / Do you sometimes have breathing difficulties on exertion? (yes/no)<br>If yes, which? <sup>a</sup> |  |
|  | • Wheeze<br>• Dyspnea<br>• Cough |  |
| Chronic cough | Has your child / Have you had cough in the last 12 months that lasted for more than 4 weeks? |  |

<sup>a</sup> Participants could report more than one symptom.

**Supplementary Table S2.**
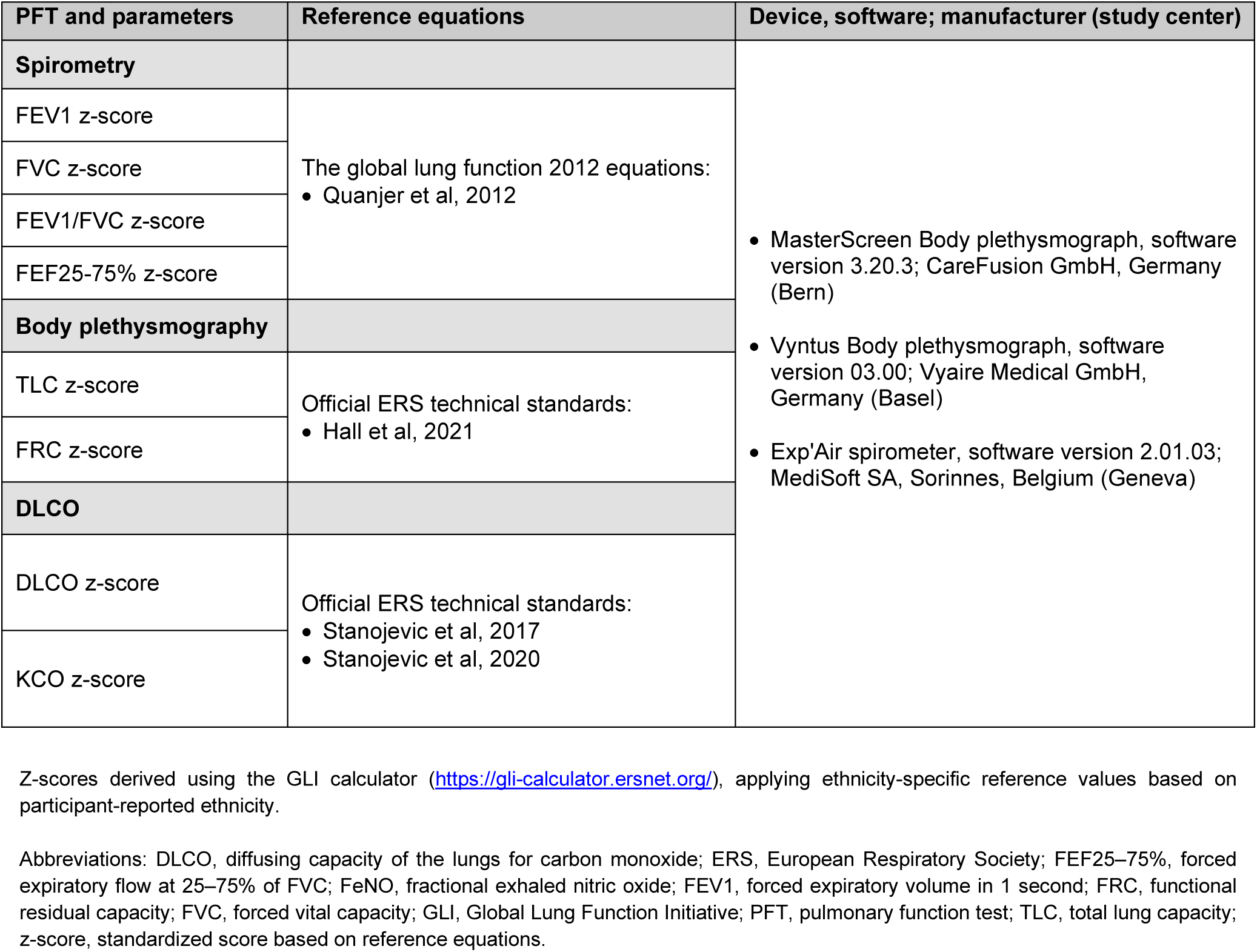
Pulmonary function test parameters, corresponding standard reference equations, and detailed specifications of measurement devices and software used at Bern, Basel, and Geneva study centers in the SCCSS FollowUp – Pulmo study.

**Supplementary Table S3.**
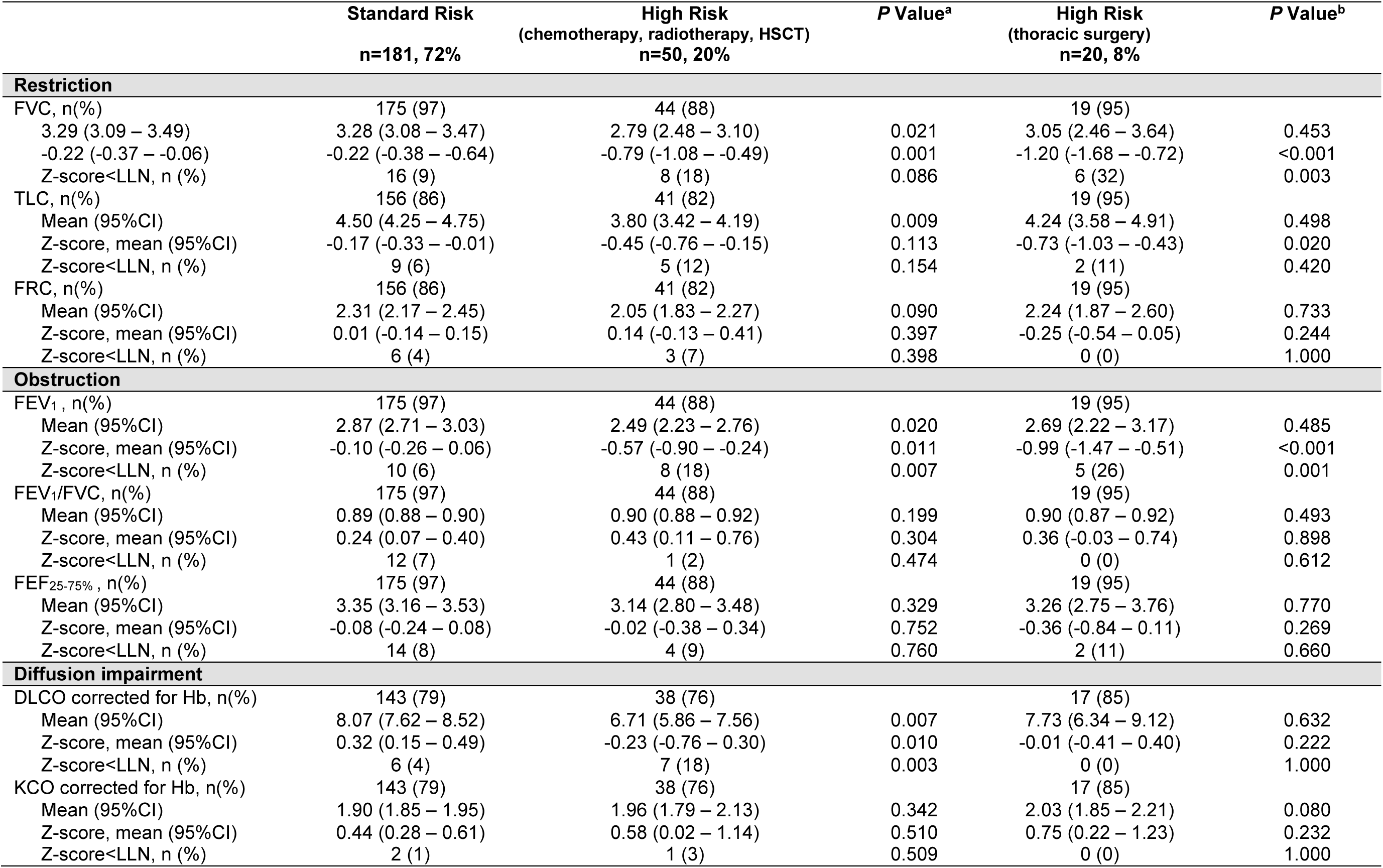
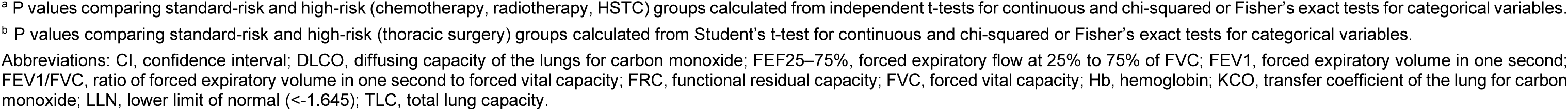
Pulmonary function parameters among childhood cancer survivors, reported by treatment-related risk groups. Parameters include measures of restriction, obstruction, and diffusion capacity, with prevalence of impairment and group comparisons.

|  | Standard Risk<br>n=181, 72% | High Risk<br>(chemotherapy, radiotherapy, HSCT)<br>n=50, 20% | P Value<sup>a</sup> | High Risk<br>(thoracic surgery)<br>n=20, 8% | P Value<sup>b</sup> |
| --- | --- | --- | --- | --- | --- |
| **Restriction** |  |  |  |  |  |
| FVC, n(%) | 175 (97) | 44 (88) |  | 19 (95) |  |
| 3.29 (3.09 – 3.49) | 3.28 (3.08 – 3.47) | 2.79 (2.48 – 3.10) | 0.021 | 3.05 (2.46 – 3.64) | 0.453 |
| -0.22 (-0.37 – -0.06) | -0.22 (-0.38 – -0.64) | -0.79 (-1.08 – -0.49) | 0.001 | -1.20 (-1.68 – -0.72) | <0.001 |
| Z-score<LLN, n (%) | 16 (9) | 8 (18) | 0.086 | 6 (32) | 0.003 |
| TLC, n(%) | 156 (86) | 41 (82) |  | 19 (95) |  |
| Mean (95%CI) | 4.50 (4.25 – 4.75) | 3.80 (3.42 – 4.19) | 0.009 | 4.24 (3.58 – 4.91) | 0.498 |
| Z-score, mean (95%CI) | -0.17 (-0.33 – -0.01) | -0.45 (-0.76 – -0.15) | 0.113 | -0.73 (-1.03 – -0.43) | 0.020 |
| Z-score<LLN, n (%) | 9 (6) | 5 (12) | 0.154 | 2 (11) | 0.420 |
| FRC, n(%) | 156 (86) | 41 (82) |  | 19 (95) |  |
| Mean (95%CI) | 2.31 (2.17 – 2.45) | 2.05 (1.83 – 2.27) | 0.090 | 2.24 (1.87 – 2.60) | 0.733 |
| Z-score, mean (95%CI) | 0.01 (-0.14 – 0.15) | 0.14 (-0.13 – 0.41) | 0.397 | -0.25 (-0.54 – 0.05) | 0.244 |
| Z-score<LLN, n (%) | 6 (4) | 3 (7) | 0.398 | 0 (0) | 1.000 |
| **Obstruction** |  |  |  |  |  |
| FEV<sub>1</sub>, n(%) | 175 (97) | 44 (88) |  | 19 (95) |  |
| Mean (95%CI) | 2.87 (2.71 – 3.03) | 2.49 (2.23 – 2.76) | 0.020 | 2.69 (2.22 – 3.17) | 0.485 |
| Z-score, mean (95%CI) | -0.10 (-0.26 – 0.06) | -0.57 (-0.90 – -0.24) | 0.011 | -0.99 (-1.47 – -0.51) | <0.001 |
| Z-score<LLN, n (%) | 10 (6) | 8 (18) | 0.007 | 5 (26) | 0.001 |
| FEV<sub>1</sub>/FVC, n(%) | 175 (97) | 44 (88) |  | 19 (95) |  |
| Mean (95%CI) | 0.89 (0.88 – 0.90) | 0.90 (0.88 – 0.92) | 0.199 | 0.90 (0.87 – 0.92) | 0.493 |
| Z-score, mean (95%CI) | 0.24 (0.07 – 0.40) | 0.43 (0.11 – 0.76) | 0.304 | 0.36 (-0.03 – 0.74) | 0.898 |
| Z-score<LLN, n (%) | 12 (7) | 1 (2) | 0.474 | 0 (0) | 0.612 |
| FEF<sub>25-75%</sub>, n(%) | 175 (97) | 44 (88) |  | 19 (95) |  |
| Mean (95%CI) | 3.35 (3.16 – 3.53) | 3.14 (2.80 – 3.48) | 0.329 | 3.26 (2.75 – 3.76) | 0.770 |
| Z-score, mean (95%CI) | -0.08 (-0.24 – 0.08) | -0.02 (-0.38 – 0.34) | 0.752 | -0.36 (-0.84 – 0.11) | 0.269 |
| Z-score<LLN, n (%) | 14 (8) | 4 (9) | 0.760 | 2 (11) | 0.660 |
| **Diffusion impairment** |  |  |  |  |  |
| DLCO corrected for Hb, n(%) | 143 (79) | 38 (76) |  | 17 (85) |  |
| Mean (95%CI) | 8.07 (7.62 – 8.52) | 6.71 (5.86 – 7.56) | 0.007 | 7.73 (6.34 – 9.12) | 0.632 |
| Z-score, mean (95%CI) | 0.32 (0.15 – 0.49) | -0.23 (-0.76 – 0.30) | 0.010 | -0.01 (-0.41 – 0.40) | 0.222 |
| Z-score<LLN, n (%) | 6 (4) | 7 (18) | 0.003 | 0 (0) | 1.000 |
| KCO corrected for Hb, n(%) | 143 (79) | 38 (76) |  | 17 (85) |  |
| Mean (95%CI) | 1.90 (1.85 – 1.95) | 1.96 (1.79 – 2.13) | 0.342 | 2.03 (1.85 – 2.21) | 0.080 |
| Z-score, mean (95%CI) | 0.44 (0.28 – 0.61) | 0.58 (0.02 – 1.14) | 0.510 | 0.75 (0.22 – 1.23) | 0.232 |
| Z-score<LLN, n (%) | 2 (1) | 1 (3) | 0.509 | 0 (0) | 1.000 |

**Supplementary Table S4.** Predictors of lung function (FEV₁ and FVC z-scores) among childhood cancer survivors. Results from multivariable linear regression models examining clinical and treatment-related exposures.

| Exposure | FEV<sub>1</sub> z-score |  |  | FVC z-score |  |  |
| --- | --- | --- | --- | --- | --- | --- |
| | $\beta$ | (95% CI) | P Value <sup>a</sup> | $\beta$ | (95% CI) | P Value <sup>a</sup> |
| Sex (ref: male) | 0.15 | (-0.11 – 0.42) | 0.258 | 0.08 | (-0.19 – 0.35) | 0.516 |
| Age at study (per year) | -0.08 | (-0.12 – -0.05) | <0.001 | -0.04 | (-0.08 – -0.01) | 0.015 |
| Time since diagnosis (per year) | 0.01 | (-0.02 – 0.05) | 0.421 | -0.02 | (-0.05 – 0.02) | 0.307 |
| Bleomycin (ref: no) | 0.43 | (-1.00 – 1.86) | 0.551 | 0.53 | (-0.88 – 1.94) | 0.463 |
| Busulfan (ref: no) | 0.27 | (-0.49 – 1.03) | 0.483 | 0.41 | (-0.34 – 1.16) | 0.284 |
| Nitrosoureas <sup>b</sup> (ref: no) | -0.71 | (-1.49 – 0.08) | 0.078 | -1.09 | (-1.87 – -0.32) | 0.006 |
| Thoracic surgery (ref: no) | -0.92 | (-1.42 – -0.41) | <0.001 | -0.97 | (-1.46 – -0.47) | <0.001 |
| Pulmotoxic radiotherapy (ref: no) | 0.01 | (-0.48 – 0.49) | 0.981 | -0.14 | (-0.62 – 0.33) | 0.551 |
| HSCT (ref: no) | -0.80 | (-1.47 – -0.13) | 0.022 | -0.84 | (-1.50 – -0.18) | 0.013 |

<sup>a</sup> P values are based on the *t*-statistics testing whether each regression coefficient differs significantly from zero in linear regression models.
<sup>b</sup> Including carmustine or lomustine.
Abbreviations: FEV<sub>1</sub>, forced expiratory volume in 1 second; FEV<sub>1</sub>/FVC, ratio of forced expiratory volume in 1 second to forced vital capacity; FVC, forced vital capacity; HSCT, haematopoietic stem cell transplantation.

**Supplementary Table S5.** Predictors of pulmonary function outcomes among childhood cancer survivors. Multivariable linear regression models showing associations of clinical characteristics and cancer treatments with FEV1/FVC, TLC z-score, DLCO z-score, FEV1 z-score, and FVC z-score, adjusted for asthma.

| Exposure | TLC z-score |  |  | FEV1/FVC |  |  | DLCO z-score |  |  |
| --- | --- | --- | --- | --- | --- | --- | --- | --- | --- |
| | $\beta$ | (95% CI) | P Value <sup>a</sup> | $\beta$ | (95% CI) | P Value <sup>a</sup> | $\beta$ | 95%CI | P Value <sup>a</sup> |
| Sex (ref: male) | 0.10 | (-0.16 – 0.37) | 0.436 | 0.03 | (0.01 – 0.05) | <0.001 | -0.22 | (-0.57 – 0.14) | 0.227 |
| Age at study (per year) | -0.02 | (-0.06 – 0.02) | 0.301 | -0.005 | (-0.007 – -0.003) | <0.001 | -0.07 | (-0.12 – -0.02) | 0.005 |
| Time since diagnosis (per year) | -0.003 | (-0.04 – 0.03) | 0.875 | 0.003 | (0.001 – 0.004) | 0.025 | 0.05 | (0.003 – 0.10) | 0.039 |
| Bleomycin (ref: no) | 0.34 | (-1.01 – 1.69) | 0.623 | -0.01 | (-0.10 – 0.07) | 0.746 | 0.60 | (-1.13 – 2.32) | 0.495 |
| Busulfan (ref: no) | 0.62 | (-0.14 – 1.38) | 0.111 | -0.02 | (-0.07 – 0.02) | 0.283 | -0.02 | (-0.99 – 0.95) | 0.969 |
| Nitrosoureas <sup>b</sup> (ref: no) | -1.38 | (-2.19 – -0.58) | 0.001 | 0.06 | (0.01 – 0.10) | 0.021 | -0.59 | (-1.71 – 0.54) | 0.303 |
| Thoracic surgery (ref: no) | -0.52 | (-1.00 – -0.04) | 0.033 | -0.001 | (-0.03 – 0.03) | 0.978 | 0.05 | (-0.64 – 0.73) | 0.893 |
| Pulmotoxic radiotherapy (ref: no) | -0.24 | (-0.71 – 0.22) | 0.306 | 0.03 | (0.002 – 0.06) | 0.034 | -0.68 | (-1.31 – -0.04) | 0.037 |
| HSCT (ref: no) | -0.32 | (-1.01 – 0.36) | 0.356 | -0.001 | (-0.04 – 0.04) | 0.975 | -0.12 | (-1.00 – 0.75) | 0.783 |
| Asthma (ref: no) | 0.16 | (-0.41 – 0.73) | 0.584 | -0.06 | (-0.09 – -0.03) | <0.001 | 0.81 | (-0.01 – 1.59) | 0.062 |
| Exposure | FEV<sub>1</sub> z-score |  |  | FVC z-score |  |  |  |  |  |
| | $\beta$ | (95% CI) | P Value <sup>a</sup> | $\beta$ | (95% CI) | P Value <sup>a</sup> | | | |
| Sex (ref: male) | 0.13 | (-0.14 – 0.39) | 0.351 | 0.08 | (-0.18 – 0.35) | 0.540 |  |  |  |
| Age at study (per year) | -0.08 | (-0.11 – -0.04) | <0.001 | -0.04 | (-0.08 – -0.01) | 0.022 |  |  |  |
| Time since diagnosis (per year) | 0.005 | (-0.03 – 0.04) | 0.793 | -0.02 | (-0.06 – 0.02) | 0.281 |  |  |  |
| Bleomycin (ref: no) | 0.40 | (-1.01 – 1.82) | 0.574 | 0.52 | (-0.89 – 1.93) | 0.468 |  |  |  |
| Busulfan (ref: no) | 0.25 | (-0.50 – 1.00) | 0.509 | 0.41 | (-0.35 – 1.16) | 0.288 |  |  |  |
| Nitrosoureas <sup>b</sup> (ref: no) | -0.69 | (-1.47 – 0.09) | 0.083 | -1.09 | (-1.87 – -0.31) | 0.006 |  |  |  |
| Thoracic surgery (ref: no) | -0.93 | (-1.42 – -0.43) | <0.001 | -0.97 | (-1.47 – -0.47) | <0.001 |  |  |  |
| Pulmotoxic radiotherapy (ref: no) | 0.06 | (-0.42 – 0.55) | 0.798 | -0.14 | (-0.62 – 0.35) | 0.579 |  |  |  |
| HSCT (ref: no) | -0.84 | (-1.51 – -0.18) | 0.013 | -0.85 | (-1.51 – -0.18) | 0.013 |  |  |  |
| Asthma (ref: no) | -0.63 | (-1.17 – -0.09) | 0.022 | -0.10 | (-0.64 – 0.44) | 0.710 |  |  |  |

<sup>a</sup> P values are based on the *t*-statistics testing whether each regression coefficient differs significantly from zero in linear regression models.
<sup>b</sup> Including carmustine and lomustine.
Abbreviations: CI, confidence interval; DLCO, diffusing capacity of the lungs for carbon monoxide; FEV<sub>1</sub>, forced expiratory volume in 1 second; FEV<sub>1</sub>/FVC, ratio of forced expiratory volume in 1 second to forced vital capacity; FVC, forced vital capacity; HSCT, haematopoietic stem cell transplantation; TLC, total lung capacity.

**Supplementary Table S6.**
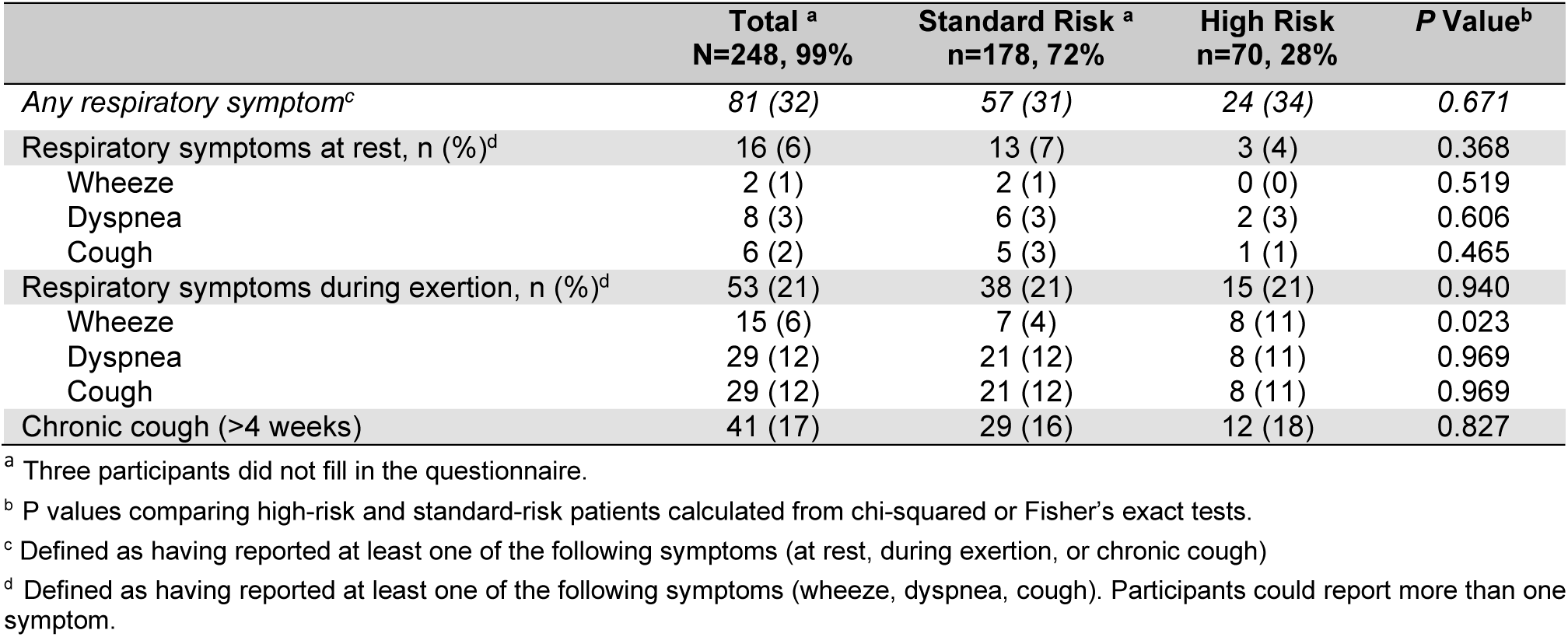
Prevalence and types of self-reported respiratory symptoms in childhood cancer survivors by treatment risk group.

**Supplementary Table S7.**
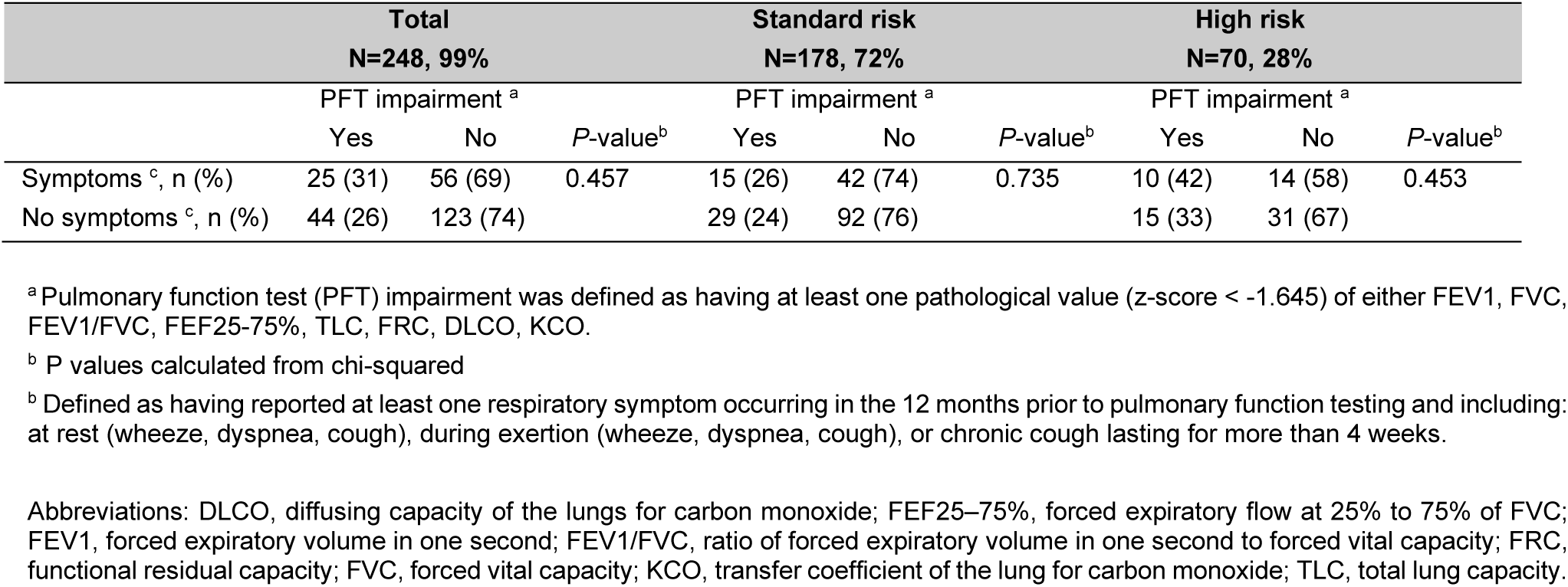
Comparison of recent respiratory symptoms by pulmonary function impairment status in childhood cancer survivors, overall and by treatment-related risk groups.

**Supplementary Table S8.**
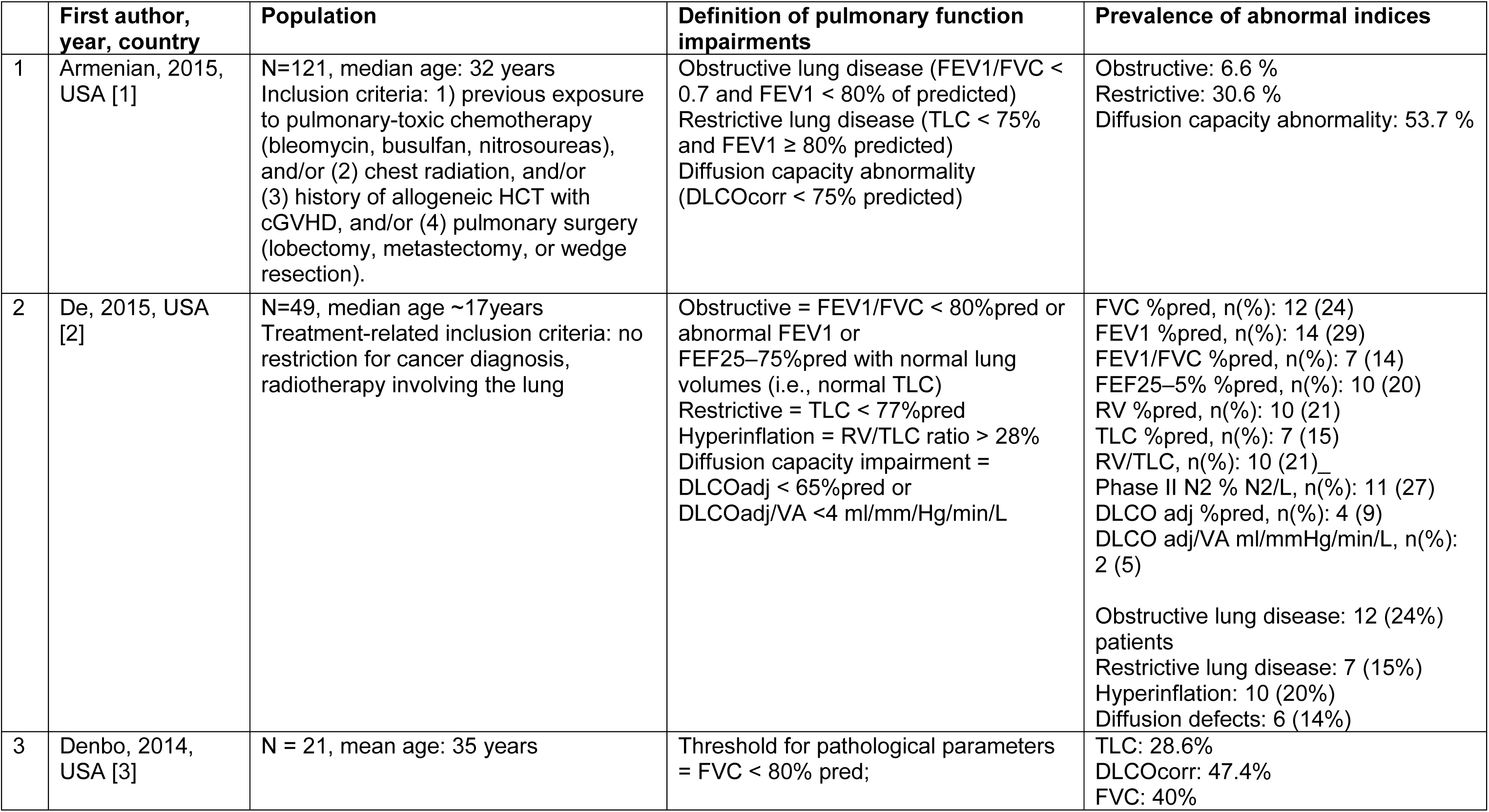
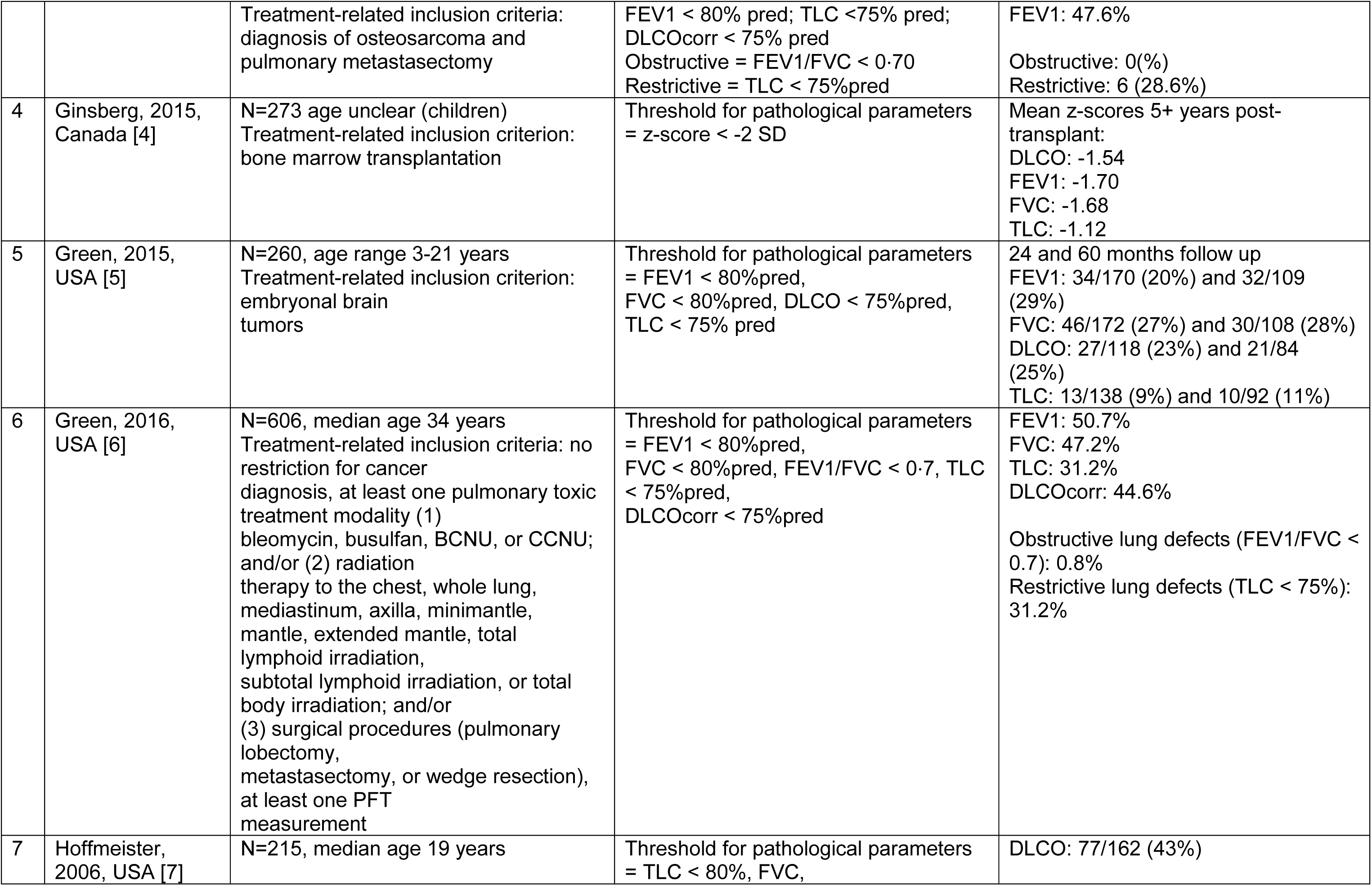
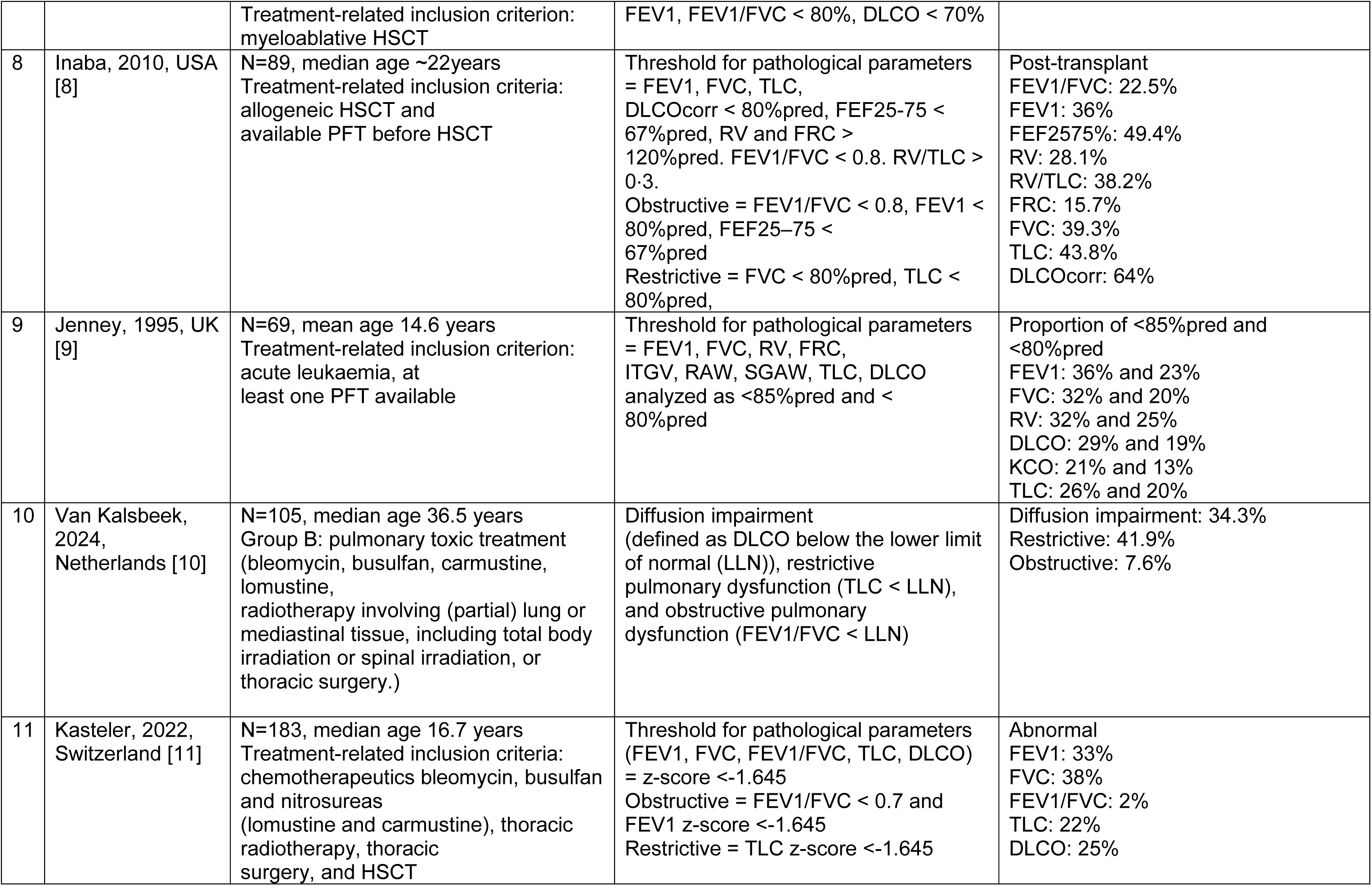
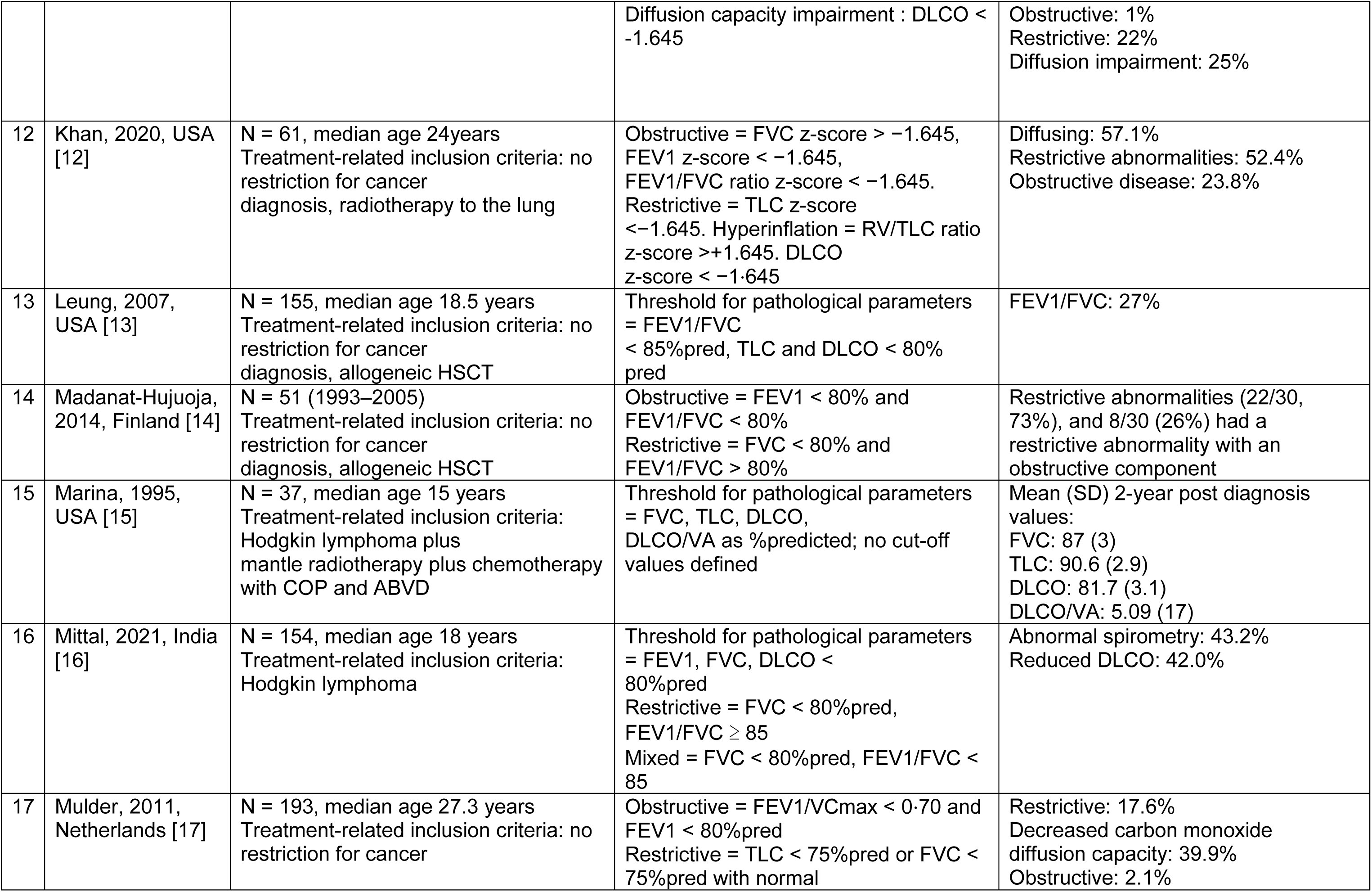
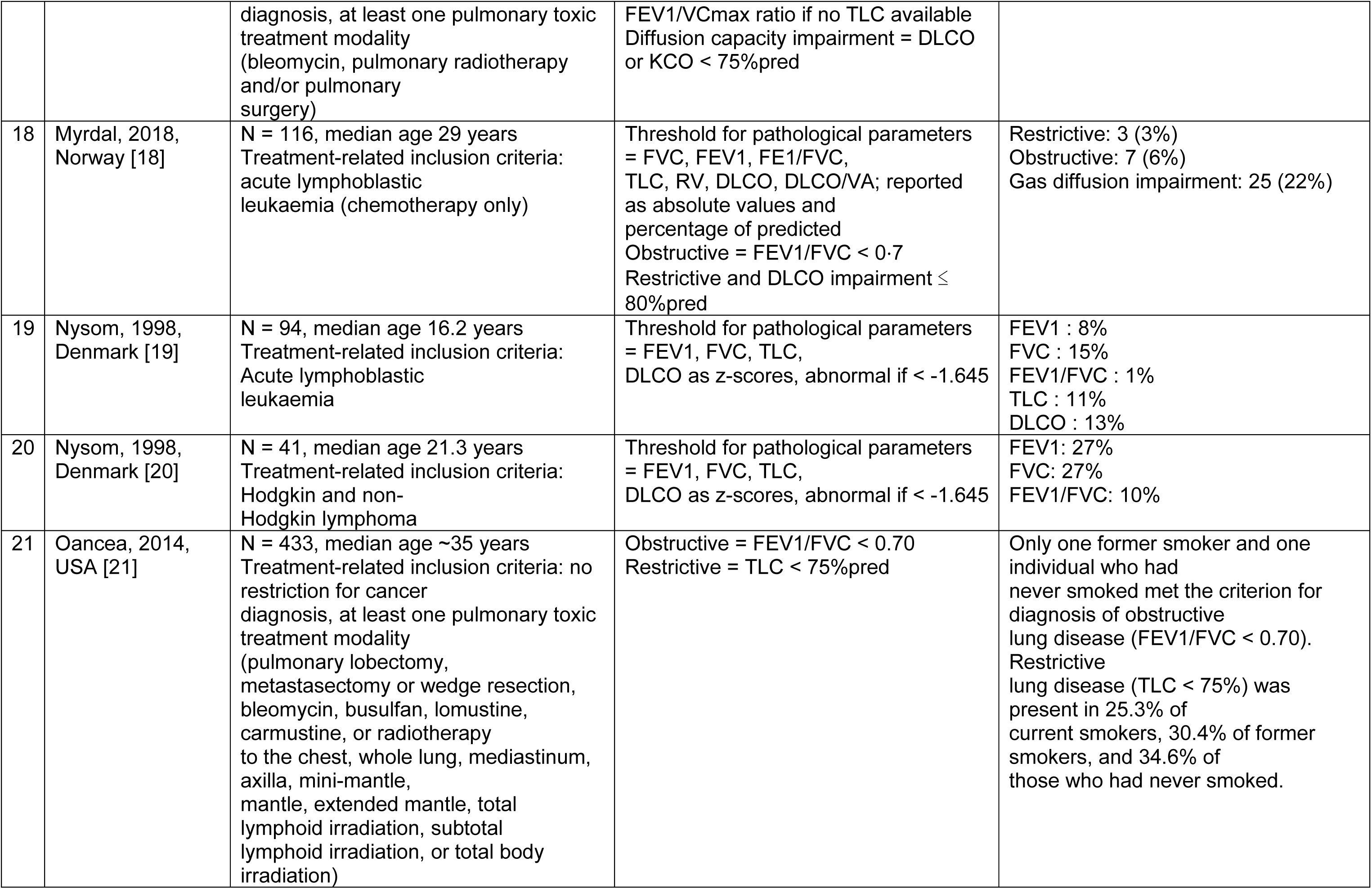
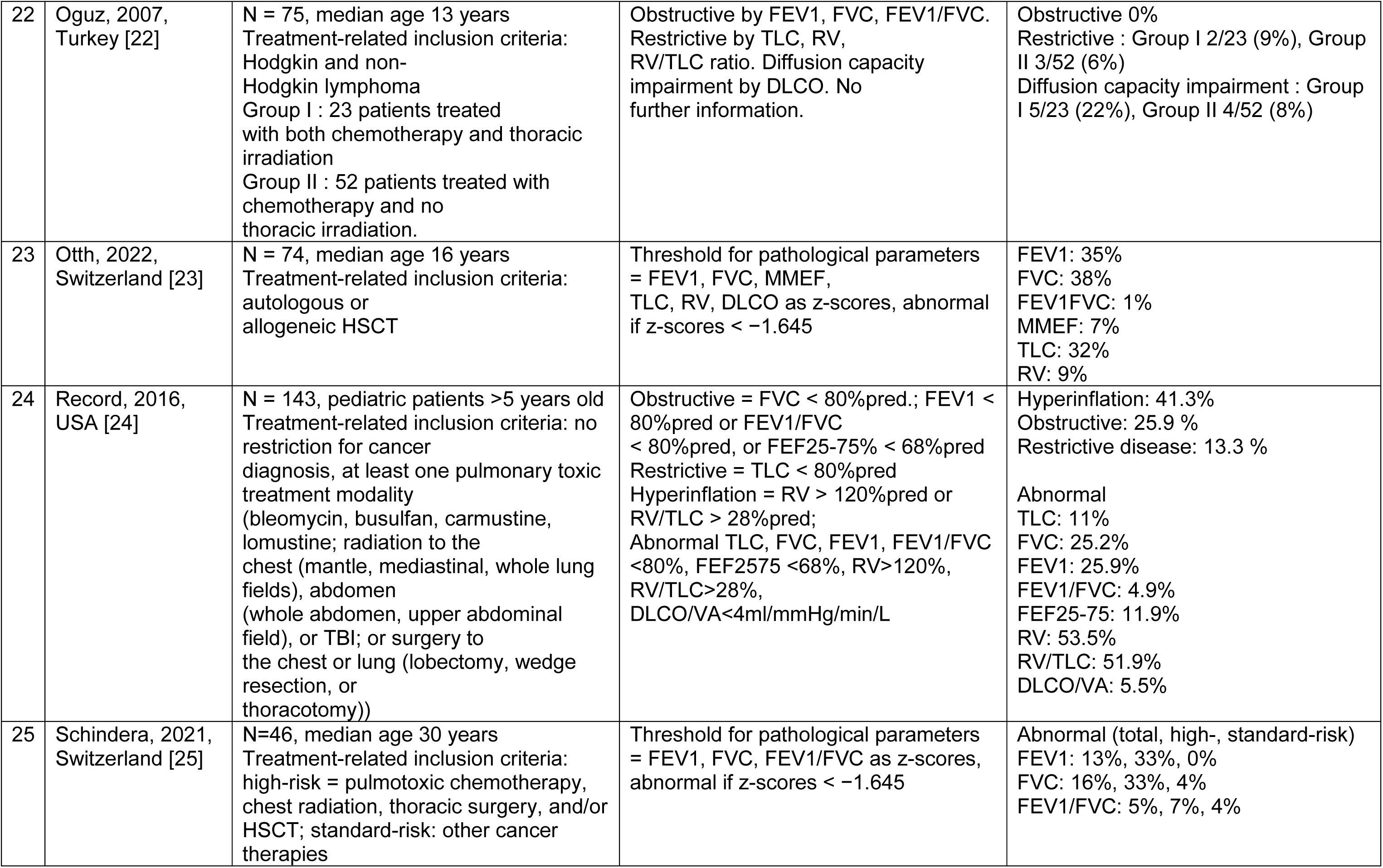
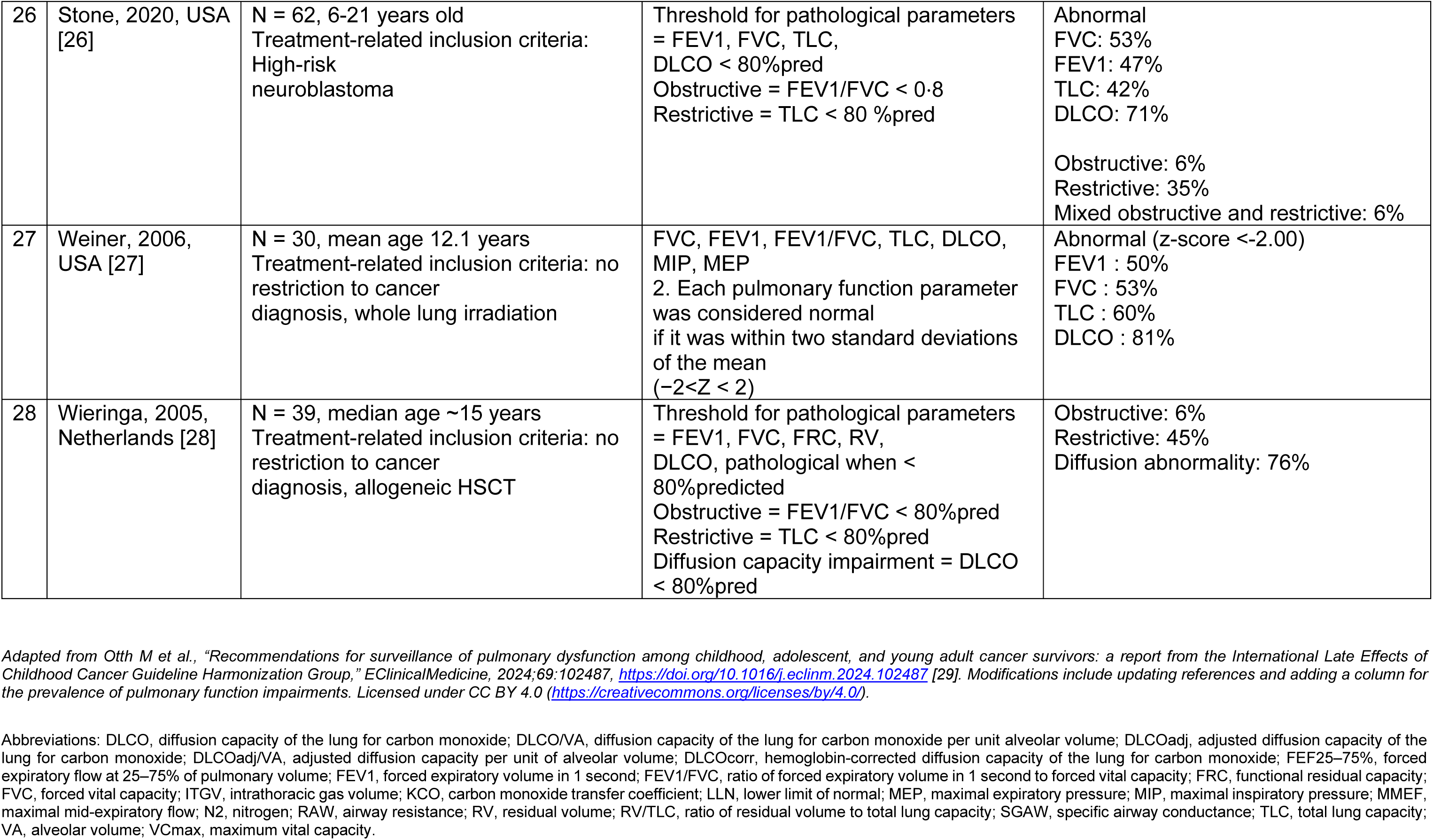
Summary of studies on pulmonary function testing in childhood cancer survivors including the prevalence of abnormal indices.

